# Short-Term Changes in Dermal Density Following Topical Application of a Progerinin-Containing Serum: An Exploratory Clinical Study

**DOI:** 10.1101/2025.09.10.25335131

**Authors:** So-mi Kang, Minju Kim, Tae-Gyun Woo, Soyoung Park, Bae-Hoon Kim, Bum-Joon Park

**Author notes:** Correspondence (Bum-Joon Park), (Bae-Hoon Kim). These authors contributed equally to this work (So-mi Kang, Minju Kim).

## Abstract

Progerin, an aberrantly processed form of lamin A, is best known as the causative protein in Hutchinson-Gilford progeria syndrome but is also detectable at low levels in normally aging human skin. Accumulation of progerin has therefore been proposed as one of the cellular processes potentially contributing to age-associated deterioration of skin homeostasis. Previous experimental studies have shown that pharmacological targeting of progerin with Progerinin can ameliorate progerin-associated cellular abnormalities and tissue phenotypes, providing a biological rationale for exploring its potential relevance to skin aging. However, whether topical formulations incorporating a progerin-targeting compound can influence measurable features of human skin has not been clinically explored. In this exploratory study, we evaluated short-term changes in skin parameters following topical application of a serum containing Progerinin, a small-molecule progerin inhibitor, together with conventional skin-conditioning ingredients, with particular attention to dermal density.

Twenty-one women aged 30-50 years participated in a 4-week, single-arm, prospective study, in which the test serum was administered once daily in the evening. Lateral canthal wrinkles, facial lifting, skin hydration, elasticity, dermal density, and skin brightness were objectively assessed at baseline and after 2 and 4 weeks of treatment, with dermal density evaluated by ultrasound imaging.

Progressive improvements from baseline were observed across the evaluated skin parameters. The most prominent change was observed in dermal density, which increased by 10.543% after 2 weeks and 23.583% after 4 weeks (p < 0.025). After 4 weeks, skin hydration increased by 18.435%, elasticity by 8.563% and skin brightness by 3.424%, whereas lateral canthal wrinkles and the facial lifting angle decreased by 5.631% and 3.234%, respectively (p < 0.05). No treatment-related adverse skin reactions were reported or identified during dermatological examination.

Four weeks of topical application of the Progerinin-containing serum was therefore associated with measurable improvements in multiple skin parameters, with a particularly pronounced and progressive increase in ultrasound-assessed dermal density. The direction of this structural skin response is consistent with the previously reported biological activity of Progerinin in progerin-driven experimental models and provides an initial translational signal supporting further investigation of progerin-targeted approaches in human skin. Because this exploratory study did not include a vehicle or placebo control and the formulation contained additional active skincare ingredients, the specific contribution of Progerinin remains to be established. Nevertheless, the magnitude and time-dependent pattern of the dermal density response, considered together with prior mechanistic and preclinical evidence for Progerinin, provide a strong rationale for its further evaluation in randomized, vehicle-controlled studies.

## INTRODUCTION

Skin aging is characterized by progressive structural and functional alterations involving both the epidermis and dermis. With increasing age, dermal fibroblast activity declines and extracellular matrix (ECM) homeostasis becomes progressively disrupted, resulting in reduced collagen content, altered elastic fibers, dermal thinning, loss of elasticity, and the development of wrinkles and skin laxity [1–5]. Environmental factors, particularly ultraviolet radiation, further accelerate these processes and contribute to photoaging [2, 3, 6–9].

Most topical anti-aging approaches have consequently focused on hydration, antioxidant activity, extracellular matrix maintenance, or stimulation of collagen synthesis [4–6, 10–15]. Increasing evidence, however, suggests that molecular processes associated with cellular aging may represent additional targets for interventions designed to maintain skin structure [4, 7, 12]. Targeting upstream cellular mechanisms associated with tissue aging may therefore offer a complementary strategy to conventional approaches that primarily address downstream structural or cosmetic manifestations.

Progerin is a truncated and permanently farnesylated form of lamin A generated by abnormal splicing of LMNA. High levels of progerin cause Hutchinson-Gilford progeria syndrome (HGPS), a rare premature-aging disorder characterized by profound abnormalities in nuclear architecture and accelerated deterioration of multiple tissues [16, 17]. Importantly, low-level progerin expression is not restricted to HGPS. Progerin has also been detected in cells and tissues from individuals without progeria, including human fibroblasts and keratinocytes, and its accumulation has been associated with cellular and tissue aging [18–23]. In human skin, progerin-positive cells have been reported to increase with chronological age, supporting the possibility that progerin-associated cellular dysfunction may contribute to age-related alterations in cutaneous homeostasis [19]. Progerin homeostasis also influenced by endogenous regulatory mechanism; notably, WRN has been identified as an intrinsic inhibitor of progerin expression [24].

These observations raise the possibility that modulation of progerin-associated cellular abnormalities may have relevance beyond rare premature-aging disorders. Progerinin is a small-molecule inhibitor developed to interfere with the pathological interaction between progerin and lamin A/C [25, 26]. Mechanistic studies targeting this interaction have demonstrated improvements in characteristic progerin-associated abnormalities, including nuclear deformation and premature cellular senescence [25, 26]. Subsequent studies with Progerinin have further demonstrated improvements in aging-associated cellular phenotypes and multiple pathological features in HGPS mouse models [26, 27]. Importantly, Progerinin has also progressed to clinical evaluation and was well tolerated in a completed early-phase clinical study, with no clinically significant safety concerns identified at the evaluated doses (ClinicalTrials.gov Identifier: NCT04512963). Of particular relevance to the present study, Progerinin treatment also improved skin-associated histological abnormalities in progeroid mice [26], providing a preclinical basis for investigation whether progerin-directed intervention may influence measurable structural properties of human skin.

Collectively, the presence of progerin in normally aging skin [18–22] and the previously demonstrated biological activity of Progerinin [25, 26, 28] provide a mechanistic and translational rationale for exploring Progerinin as a potential component of interventions aimed at maintaining age-associated skin structure. Whether a Progerinin-containing topical formulation can influence measurable properties of human skin, however, remains unknown.

We therefore conducted an exploratory 4-week human application study using a topical serum containing Progerinin together with conventional skincare ingredients. Multiple objective skin parameters were evaluated longitudinally, including wrinkles, facial lifting, hydration, elasticity, brightness, and ultrasound-assessed dermal density.

Because the study was designed as a single-arm exploratory investigation without a vehicle-control group, it was not intended to establish the independent efficacy of Progerinin. Rather, the objective was to determine whether topical application of a Progerinin-containing formulation could generate a measurable clinical response in human skin and whether the pattern of this response would support further controlled investigation of Progerinin. Particular attention was given to dermal density because it provides an objective, structure-related measure of the dermis and showed the largest relative change during the study.

## MATERIALS AND METHODS

### 1. Test Product

The general information of the test product is shown in **Table 1**.

### 2. Distribution of Test Product

The test product was provided by the sponsor, PRGS & Tech Co., Ltd., and was used by the test subjects with labels attached, which specified the test number, subject number, product name, manufacturer, storage method, etc.

### 3. Usage Instructions for the test product

After cleansing, take an appropriate amount on the fingertips and gently apply it to the required areas, tapping gently to help absorption. Additionally, during the human application test period, the use of any cosmetics containing active ingredients that could affect the test results (e.g., functional cosmetics for whitening, wrinkle improvement, etc.) other than the test product provided by this center was strictly prohibited. The frequency of use of test product was 4 weeks (once a day in the evening). The test product was stored in a sealed condition at room temperature.

### 4. Ingredients of the skin serum

The information on the ingredients is shown in **Table 2**.

### 5. Selection Criteria of Test Subjects

The following criteria were established for participant selection in the clinical trial evaluating the efficacy of the skin serum: Adult women aged 30 to 50 years who exhibit visible eye wrinkles were considered suitable candidates for the study. Participants were required to be fully informed about the trial details by the principal investigator or an authorized delegate. They had to voluntarily agree to participate by signing a written informed consent form. Only healthy individuals without any infectious skin diseases or acute or chronic physical conditions were included in the trial to ensure unbiased results and participant safety. Participants needed to be available for follow-up visits and assessments throughout the trial period to facilitate accurate data collection and evaluation of results. These criteria were carefully designed to ensure the selection of an appropriate study population and to maintain the validity and reliability of the clinical trial outcomes.

### 6. Exclusion Criteria of Test Subjects

Participants who met any of the following criteria were excluded from the clinical trial to ensure the safety and reliability of the study: Women who were pregnant, breastfeeding, or had the potential to become pregnant during the trial were excluded to avoid risks to both the participant and a potential fetus. Individuals who declined to participate or did not provide signed informed consent were not eligible. Participants with psychiatric disorders that could affect compliance or understanding of the trial procedures were excluded. Individuals who had undergone immunosuppressive therapy within 3 months prior to the trial were excluded. Individuals who had received systemic steroids or phototherapy within 1 month prior to the trial were excluded. Those with lesions in the test area that could interfere with accurate measurement were excluded. Individuals with atopic skin or severe reactions or allergies to cosmetics, medications, or sun exposure were excluded. Participants who had undergone skin peeling or cosmetic treatments within 3 months prior to the trial were excluded to avoid confounding effects. If the principal investigator or the responsible person deemed participation unfeasible due to the above factors or other considerations, the individual was excluded. These exclusion criteria were meticulously designed to minimize risks, ensure participant safety, and maintain the integrity of the study results.

### 7. Criteria for Discontinuation and Withdrawal

Test subjects who initially agreed to participate in the trial but meet any of the following criteria will be withdrawn from the human application test: 1) If the test subject expresses a refusal to continue participation. 2) If the test subject experiences a serious adverse reaction or develops abnormal reactions, such as erythema, at the test site, and requests to stop the trial. 3) If the test product causes hypersensitivity reactions. 4) If use is discontinued due to other health conditions. 5) If there are any other unavoidable circumstances. 6) If the test subject does not comply with the prescribed trial procedures. 7) If the test subject fails to be followed up during the observation period

### 8. Number of Test Subjects

A total of 23 individuals were selected for the initial screening for this human application test, and all of them met the selection criteria and participated in the trial. Out of the 23, 2 individuals withdrew midway, so 21 individuals completed the trial.

### 9. Test Subject Visit Schedule

Visit 1 (Screening, Test Subject Selection, and Skin Measurements): After explaining the study, test subjects who agreed to participate and signed the consent form underwent demographic surveys, screening for inclusion/exclusion criteria, and medical history investigations. The method for using the test product was explained, and the test product was then applied. Before using the test product, measurements were taken for eye wrinkles, facial lifting, skin moisturizing, skin elasticity, skin density, and skin tone (brightness). Visit 2 (Skin Measurements): After Visit 1, any adverse reactions and concurrent treatments were investigated. Two weeks after using the test product, measurements were taken again for eye wrinkles, facial lifting, skin moisturizing, skin elasticity, skin density, and skin tone (brightness), along with an evaluation of skin conditions. Visit 3 (Skin Measurements and Questionnaire): After Visit 2, any adverse reactions and concurrent treatments were again reviewed. Four weeks after using the test product, measurements were taken for eye wrinkles, facial lifting, skin moisturizing, skin elasticity, skin density, and skin tone (brightness), along with an evaluation of skin conditions. After the trial was completed, effectiveness and preference surveys were conducted.

### 10. Evaluation Areas and Measurement Methods

For device-based evaluations, the test subjects were allowed to rest in a controlled environment (temperature: 20–25°C, humidity: 40–60%) for 30 minutes to allow the skin surface temperature and humidity to adapt to the environment. During this resting period, water intake was restricted. An investigator conducted the objective measurements, and the same area was measured for each test. 1) Eye Wrinkle Measurement: Eye wrinkles were measured using the 3D skin imaging device, Antera 3D CS (Miravex Ltd., Ireland). The same area around the eyes was photographed before and after using the test product. Specific regions of the eye wrinkle area were selected from the stored images, and the Indentation Index (A.U.) value within the specified area was analyzed. A decrease in the Indentation Index indicates an improvement in skin wrinkles. 2) Facial Lifting Measurement: Facial lifting was measured using the F-ray device (BEYOUNG Ltd., Korea), which creates a contour map of the skin. The images captured by the device were analyzed using Image-ProⓇ Plus (Media Cybernetics, USA). The measurements were taken on the same cheek area before and after using the test product, and the analysis was based on the curves formed near the test area. 3) Facial (cheek) lifting measurement: A circle formed near the cheekbone was used as the reference. A straight line was drawn from the center of the circle to the corner of the mouth, and the angle between this line and a horizontal line was measured (**Supplementary Figure 1**). A smaller angle between the two lines indicates a lifting effect. 4) Skin Moisturizing Measurement: Skin moisturizing was evaluated using the Corneometer CM825 (Courage+Khazaka Electronic GmbH, Germany). The same right cheek area was measured before and after using the test product. During measurement, the Corneometer probe was placed in contact with the skin, and the test was performed three times using the sensor. The average value of these measurements was used as the hydration assessment data. The Corneometer operates on the principle that skin, as a basic insulator, allows electricity to pass more easily when it contains more moisture. It measures the capacitance at the contact point of the probe. Thus, the moisture content and capacitance are directly proportional, meaning that a higher measurement value indicates a higher moisture content. The unit of measurement is an arbitrary unit (A.U.), which is a unit constant. 5) Skin Elasticity Measurement: Skin elasticity was measured using the Cutometer MPA580 (Courage-Khazaka Electronic GmbH, Germany). The same right cheek area was measured before and after using the test product, and the R2 (recovery force) value was used as the skin elasticity evaluation data. The R2 value represents overall elasticity, and the closer the value is to 1, the more elastic the skin is. 6) Skin Density Measurement: Skin density was measured using an ultrasound imaging device, the Skin Scanner DUB-USB (TPM Taberna Pro Medicum, Germany). The same left eye area was imaged before and after using the test product. The unit of measurement was density (%). An increase in the value indicates an improvement in skin density. 7) Skin Tone (Brightness) Measurement: Skin tone brightness was measured using the VISIA CR (Canfield Imaging Systems, USA) before and after using the test product. The same facial area of the test subject was imaged. The captured images were analyzed using Image-ProⓇ Plus. A specific area in the image was selected, and the RGB values of that area were obtained and then converted to CIELab values. The L* value (lightness), which indicates skin brightness, was used to assess skin tone. A higher L* value indicates an improvement in brightness. 8) Efficacy Questionnaire (Global Assessment of Efficacy): After using the test product, test subjects were asked to respond to a questionnaire regarding the measurement items using a 5-point scale: Very Good (4), Good (3), Average (2), Poor (1), Very Poor (0). The researcher calculated the percentage of test subjects for each response and assessed the efficacy of the test product based on these results. 9) Product Preference Questionnaire: After using the test product, test subjects were asked to fill out a questionnaire about their experience with the product. The evaluation criteria included skin moisture, smoothness, spreadability, absorbency, fragrance, and overall feel. The subjects rated each item using a 5-point scale: Very Good (4), Good (3), Average (2), Poor (1), Very Poor (0). 10) Safety Evaluation: The safety of the test product was assessed by reviewing all reported adverse reactions during the trial period for all test subjects who used the product. The incidence rate of adverse reactions was calculated and used as safety evaluation data for the product. 11) Adverse Reaction Evaluation: Any abnormal skin symptoms that occurred during the use of the test product were monitored through a questionnaire throughout the trial period to assess the occurrence and severity of the symptoms. Test subjects were instructed to immediately report any unusual symptoms to the study coordinator. If adverse reactions were reported, the study coordinator informed the principal investigator. The principal investigator then assessed the severity of the symptoms, determined the potential association with the test product, and decided on appropriate actions, including whether the subject could continue participating in the trial.

### 11. Evaluation Criteria

1) Primary Efficacy Evaluation Variables: The primary efficacy evaluation variables for the test product were based on the measurements taken before and after the use of the test product, including eye wrinkles, facial lifting, skin moisturizing, skin elasticity, skin density, and skin tone brightness. 2) Secondary Efficacy Evaluation Variables: The secondary efficacy evaluation variables for the test product were based on the results of the efficacy assessment questionnaire regarding skin improvement after using the test product.

### 12. Statistical Analysis

To determine the significance of the changes in measurements before and after using the test product, statistical analysis was performed using SPSS 19.0 software. Statistical significance was determined at a 95% confidence interval with a p-value < 0.05. The p-values were rounded to three decimal places. Continuous variables were summarized as means and standard deviations, while categorical variables were summarized as frequencies and percentages. For data with three or more repeated measurements, normality tests were performed, and parametric methods such as repeated measures ANOVA were used, followed by post-hoc tests using the Bonferroni method. For non-parametric methods, the Friedman test was used, and paired comparisons were performed using the Wilcoxon signed-rank test, with post-hoc significance adjustments using the Bonferroni method.

### 13. Safety Protection of Test Subjects

This human application test was conducted in accordance with the Declaration of Helsinki, respecting human dignity and rights, and ensuring that no harm or disadvantage would come to the test subjects. The study coordinator confirmed the health status of each test subject before enrollment to ensure they were fit to participate in the study. The study coordinator was thoroughly familiar with the test product and made every effort to ensure the safety of the test subjects.

### 14. Informed Consent and Consent Explanation

Before the study began, the study principal investigator and the study coordinator provided detailed explanations of the study, including the inclusion and exclusion criteria, to test subjects who met all the requirements. Test subjects (or their guardians) were given ample opportunity to understand all aspects of the study and any potential outcomes. The information provided to the test subjects was documented, and the study principal investigator confirmed their consent by signing the informed consent form.

### 15. Confidentiality

The names of all test subjects involved in the study were kept confidential. The signed informed consent forms were stored by the researchers, and the study coordinator or monitor managed a separate list containing the test subject numbers, initials, and names, which was used for future records and evaluation.

### 16. Other Protective Measures for Test Subjects

The PNK Skin Clinical Research Center had the necessary facilities and expert personnel in place to ensure the proper conduct of the study, as outlined in the study protocol, and to prioritize the safety of the test subjects. Researchers were informed in advance about the potential adverse reactions and precautions stated in the study protocol, and any adverse events occurring during the study were addressed appropriately and reported to the sponsor. In the case of any direct or indirect harm resulting from participation in this human application test, the study principal investigator or the study coordinator would take all necessary actions to treat the injury. If any adverse reactions or damage occurred as a result of the test product or during the treatment of adverse reactions, the sponsor (PRGS & Tech Co., Ltd.) would provide compensation for the damage caused directly by the test product. However, hospitalization fees, examination costs, and consultation fees unrelated to the study would be the responsibility of the test subject.

## RESULTS

### Participant characteristics and treatment compliance

Twenty-three participants were initially enrolled, of whom two discontinued participation (**Table 3**). Twenty-one participants completed the 4-week study and were included in the efficacy analysis. All participants were female, with a mean age of 43.3 years (**Table 4**).

Among the 21 participants, six were classified as having dry skin, twelve as moderately dry skin, and three as having normal skin (**Table 5**). None reported dermatological conditions considered likely to interfere with evaluation of the test product (**Table 6**).

Overall treatment compliance was 99.320%, and no participant completing the study demonstrated compliance below 80% (**Table 7**).

### Progressive increase in dermal density during topical treatment

Among the objective parameters evaluated, dermal density exhibited the largest relative change during the 4-week treatment period (**Figure 1A and 1B**).

**Figure 1.**
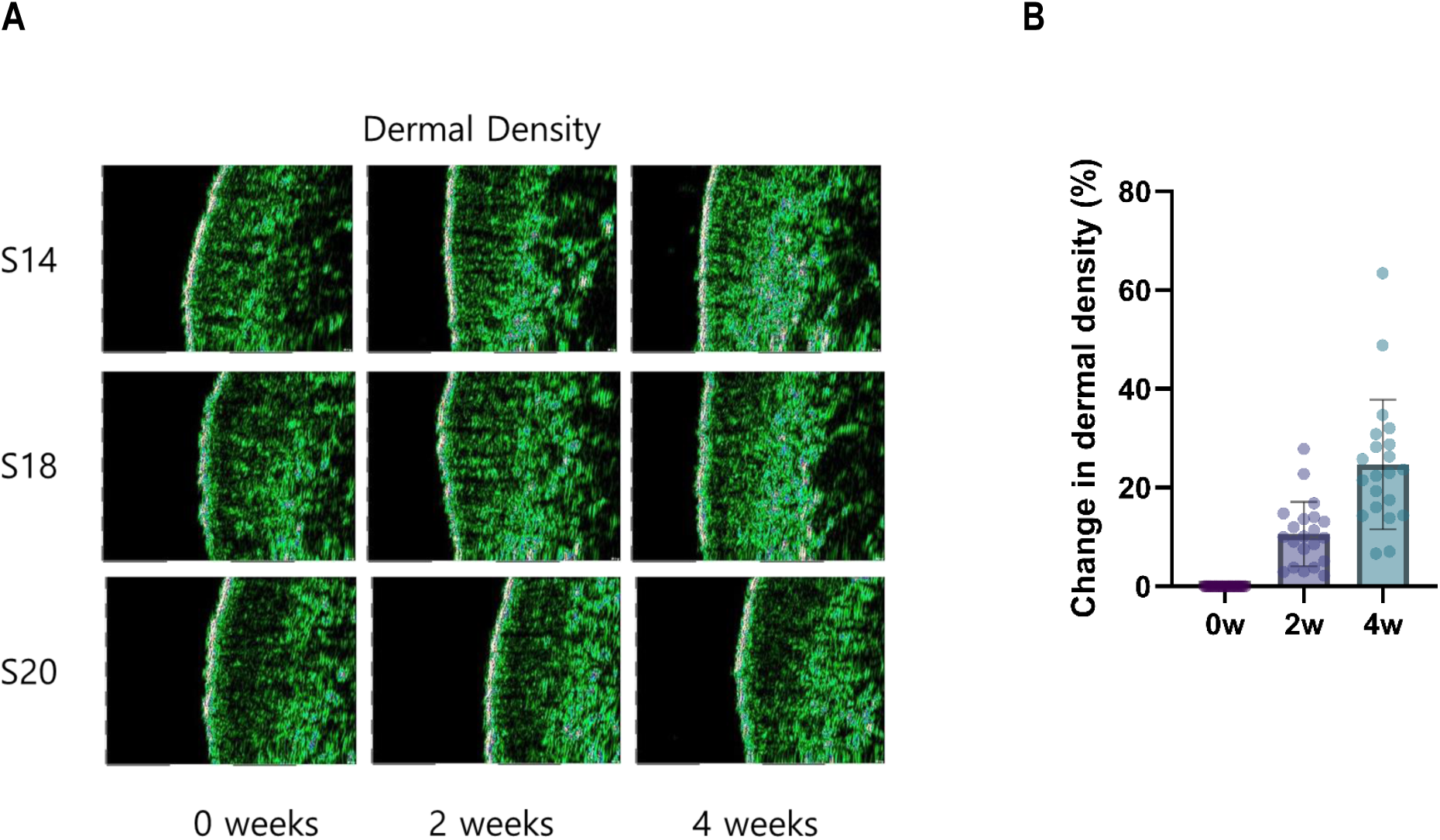
Changes in ultrasound-assessed dermal density during 4 weeks of topicalapplication of the Progerinin-containing serum. (A) Representative ultrasound images of dermal density obtained at baseline (0 weeks) and after 2 and 4 weeks of treatment from three participants (S14, S18, and S20). The ultrasound images show a progressive increase in dermal echogenic density during the treatment period. (B) Quantitative changes in dermal density relative to baseline at 0, 2, and 4 weeks. Individual data points represent individual participants. Dermal density increased progressively over the 4-week treatment period, reaching 10.543% at 2 weeks and 23.583% at 4 weeks relative to baseline. Statistical analyses were performed using the original measurement values by the Friedman test followed by Wilcoxon signed-rank tests with Bonferroni correction. Dermal density was significantly increased from baseline at both Weeks 2 and 4 (p < 0.025).

Compared with baseline, ultrasound-assessed dermal density increased by 10.543% after 2 weeks and by 23.583% after 4 weeks (p < 0.025) (**Figure 1B and Table 8**). At Week4, individual increases in dermal density ranged from 6.630% to 63.433% relative to baseline (**Figure 1B**).

Importantly, the magnitude of the change increased between Weeks 2 and 4, demonstrating a progressive temporal pattern during continued product application.

Representative ultrasound images were consistent with the quantitative measurements and showed increased dermal echogenic density following treatment.

These findings identify dermal density as the most prominent structural parameter altered during the 4-week application period.

### Changes in other skin parameters

Improvements were also observed in several additional skin parameters.

Skin hydration increased by 10.448% after 2 weeks and by 18.435% after 4 weeks relative to baseline (**Figure 2A and Table 9**). Skin elasticity increased by 4.500% and 8.563% at Weeks 2 and 4, respectively (**Figure 2B and Table 10**).

**Figure 2.**
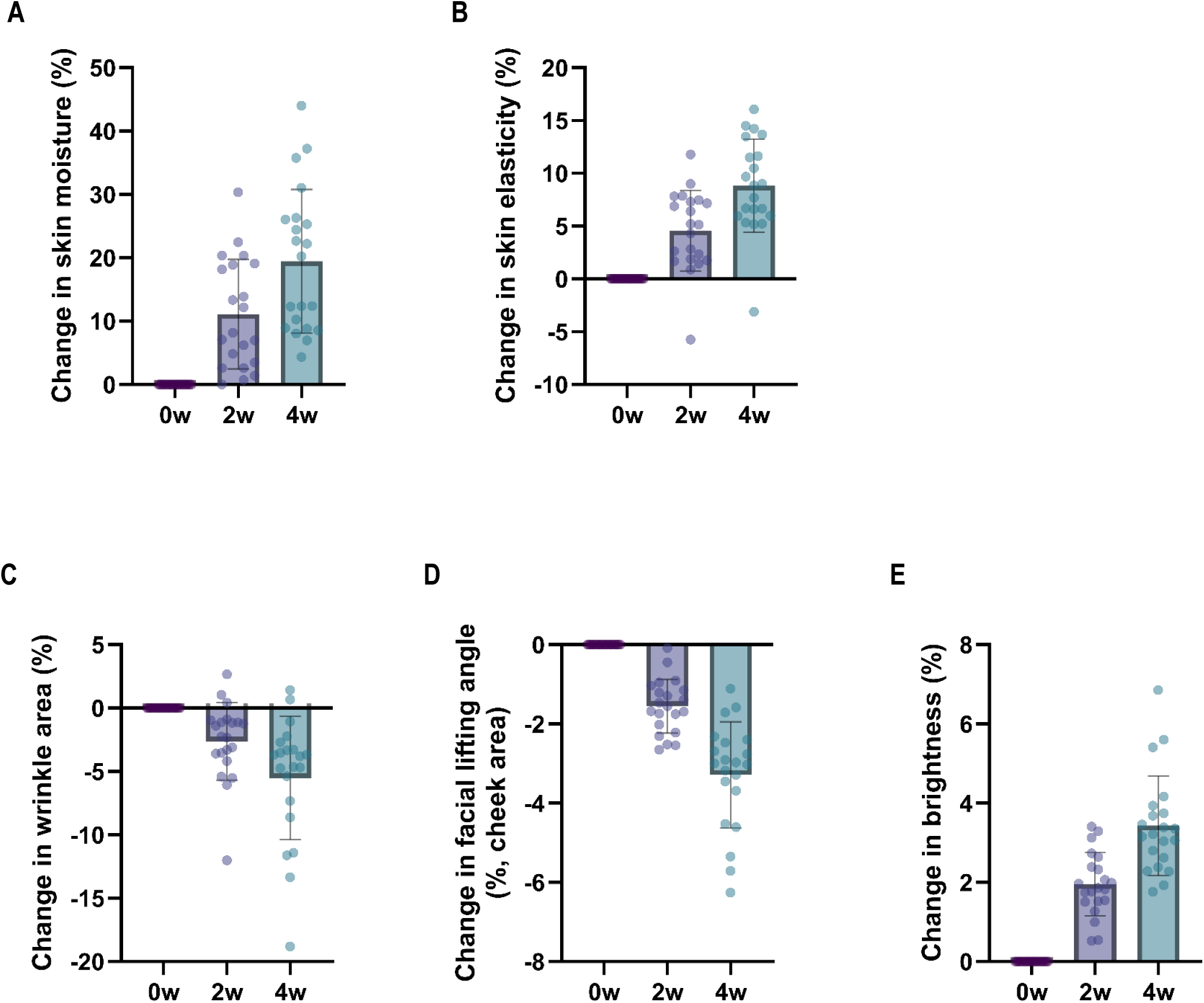
Changes in multiple skin parameters during 4 weeks of topical application of the Progerinin-containing serum. Changes from baseline were assessed after 2 and 4 weeks of treatment for (A) skin hydration, (B) skin elasticity, (C) crow’s feet wrinkles, (D) facial lifting angle, and (E) skin brightness. Skin hydration, elasticity, and brightness progressively increased during the treatment period, whereas crow’s feet wrinkles and facial lifting angle progressively decreased, consistent with improvements in these parameters. Changes are expressed as percentages relative to baseline. Statistical analyses were performed using the original measurement values by repeated-measures ANOVA followed by Bonferroni-corrected post hoc comparisons. All parameters showed statistically significant changes from baseline at both Weeks 2 and 4 (p < 0.05).

Crow’s feet wrinkles decreased by 2.628% after 2 weeks and by 5.631% after 4 weeks (**Figure 2C and Table 11**). The facial lifting angle decreased by 1.515% after 2 weeks and by 3.234% after 4 weeks, consistent with improvement in the lifting measurement (**Figure 2D and Table 12**).

Skin brightness increased by 1.951% after 2 weeks and by 3.424% after 4 weeks (**Figure 2E and Table 13**).

Together, these measurements indicate progressive changes in multiple skin characteristics during the treatment period, although the magnitude of change differed substantially among individual endpoints.

### Participant-reported outcomes and safety

Participant-reported assessments were generally consistent with the instrument-based measurements. Most participants rated improvements in wrinkles, facial lifting, hydration, elasticity, dermal density, and brightness as average or better (**Table 14 and Table 15**).

The test serum was well tolerated throughout the study. No specific adverse skin reactions were reported by participants, and dermatological examinations identified no clinically significant abnormalities related to product application (**Table 16 and Table 17**).

## DISCUSSION

The present exploratory study evaluated short-term changes in human skin following 4 weeks of daily application of a topical serum containing Progerinin together with commonly used skincare ingredients. Improvements from baseline were detected across several objectively measured parameters, including wrinkles, facial lifting, hydration, elasticity, dermal density, and brightness. Among these outcomes, the most prominent change was observed in ultrasound-assessed dermal density, which increased by 10.543% after 2 weeks and by 23.583% after 4 weeks (**Figure 1**). This substantial and progressive structural response is of particular interest in the context of the previously demonstrated biological activity of Progerinin in progerin-associated cellular and tissue abnormalities.

Dermal density is an established ultrasound-derived parameter in cosmetic and dermatologic research [29] and provides information related to the echogenic characteristics and structural organization of the dermis. Because dermal echogenicity is influenced by tissue composition and acoustic interfaces within the extracellular matrix, including collagen-rich structures, a higher ultrasound-derived density can be consistent with changes in dermal structural properties [30–32].

Several previous topical cosmetic studies have demonstrated that measurable increases in ultrasound-assessed dermal density or echogenicity can occur within relatively short treatment periods. For example, a 4-week study of a cosmetic formulation containing three-dimensional cultured adipose tissue-derived mesenchymal stem-cell conditioned medium reported increases in dermal density of 6.97% after 2 weeks and 12.53% after 4 weeks [33]. Similarly, a clinical study of hyaluronic acid formulation using the same type of ultrasound system employed in the present study reported an approximately 16.4% relative increase in skin density after 4 weeks [34]. Studies using other high-frequency ultrasound systems have likewise reported measurable short-term increases in dermal density or echogenicity following treatment with peptide- or dihydromyricetin-containing serums [35, 36]. Collectively, these findings indicate that ultrasound-derived dermal parameters are responsive to topical interventions over relatively short treatment periods and provide a useful context for interpreting the magnitude of change observed in the present study.

Evidence from a randomized split-face study further demonstrates that substantial increases in ultrasound-derived dermal density can occur following topical treatment when compared with a control formulation [37]. In that study, a crosslinked hyaluronic acid serum increased dermal density from 17.1% at baseline to 22.2% after 28 days and 25.6% after 56 days, whereas the corresponding base-cream-treated side showed only minor changes. The increase on the serum-treated side corresponded to an approximately 29.8% relative increase after 28 days [37]. These controlled findings demonstrate that increases exceeding 20% in ultrasound-derived dermal density can occur within approximately 4 weeks of topical treatment and also underscore the value of an appropriate control in evaluating formulation-associated effects.

Within this context, the 23.583% increase in dermal density observed after 4 weeks represents a substantial short-term response, with a magnitude comparable to pronounced improvement reported for the crosslinked hyaluronic acid serum in the controlled study described above. Individual responses ranged from 6.630% to 63.433% above baseline, indicating considerable inter-individual variability while also demonstrating that pronounced dermal responses were achievable in some participants. Together with the progressive increase in mean dermal density from 10.543% at Week 2 to 23.583% at Week 4, these findings support further investigation of whether a longer treatment duration or optimized topical delivery could enhance the magnitude and consistency of the response. Although differences in study design, formulation, participant characteristics, and ultrasound methodology preclude direct quantitative comparison, the magnitude and progressive pattern of the response raise the possibility that Progerinin may have contributed to the structural changes observed with formulation. This possibility is biologically relevant in the context of photoaging, as UV exposure has been reported to induce progerin expression in skin cells, including dermal fibroblasts [20–22], linking an established environmental driver of skin aging to the molecular target of Progerinin. Thus, incorporating a progerin-targeting compound into a conventional topical formulation may provide an additional mechanism to address progerin-associated cellular alterations that accompany intrinsic and UV-related skin aging.

The formulation evaluated in the present study contained Progerinin together with established skincare ingredients, including sodium hyaluronate, collagen, adenosine, niacinamide, and magnesium ascorbyl phosphate, which may independently or collectively contribute to the observed improvements [10, 13–15, 27, 31, 38]. Nevertheless, Progerinin differs from these conventional skincare ingredients in that it targets progerin-associated cellular abnormalities, a mechanism of particular relevance given the accumulation of progerin in aging skin and its induction by UV exposure [20–22]. Together with previous mechanistic and preclinical evidence showing that Progerinin ameliorates progerin-associated cellular and tissue abnormalities [25, 26], the substantial and progressive dermal response observed here supports further evaluation of Progerinin as a potential contributor to the formulation-associated effects. The small, all-female cohort and absence of a randomized control remain important limitations, and a controlled study comparing otherwise identical formulations with and without Progerinin will be required to determine its specific contribution.

The biological rationale for a potential contribution of Progerinin is supported by the presence of progerin in normally aging human skin, where progerin-positive cells increase with chronological age [18, 19, 23]. Progerin accumulation is associated with abnormalities in nuclear architecture, chromatin organization, cellular stress responses, and senescence [17, 18, 20, 25], which may impair dermal fibroblast function and extracellular matrix homeostasis [4, 30–32, 39, 40]. Progerinin was specifically developed to target this progerin-associated pathology by interfering with the pathological progerin-lamin A/C interaction, and previous mechanistic and preclinical studies demonstrated improvements in nuclear abnormalities, premature senescence-associated phenotypes, and multiple tissue-level abnormalities in HGPS models [25, 26, 28]. Of particular relevance to the present study, Progerinin treatment also improved skin-associated histological features, including enrichment of dermal connective tissue in HGPS mice [26], providing a biologically plausible link between progerin-targeted intervention and the structural skin response observed in the present study.

Cutaneous progerin levels were not measured before or after treatment, and the present study therefore cannot determine whether topical application altered progerin abundance or activity in human skin. Similarly, extracellular matrix components were not directly measured. Accordingly, the mechanistic relationship among topical Progerinin exposure, cutaneous progerin biology, and the observed dermal density response remains to be established. Direct molecular assessment in future studies would provide an important opportunity to determine whether the observed structural response is accompanied by changes in cutaneous progerin and downstream markers of dermal homeostasis.

The present findings therefore extend the investigation of Progerinin from progerin-driven experimental systems to human skin and provide initial proof-of-concept evidence that a Progerinin-containing topical formulation can produce a substantial and progressive dermal structural response. Future randomized and controlled studies integrating ultrasound imaging with direct molecular assessment of cutaneous progerin and extracellular matrix markers will be important for determining the specific contribution of Progerinin and whether the observed clinical response is associated with modulation of progerin-related skin biology.

## CONCLUSION

In this exploratory single-arm study, 4 weeks of daily application of a Progerinin-containing topical serum was associated with improvements in multiple objectively measured skin parameters. The most prominent finding was a progressive increase in ultrasound-assessed dermal density, reaching 23.583% above baseline after 4 weeks. The magnitude and progressive pattern of this response, together with the established biological activity of Progerinin in progerin-associated cellular and tissue abnormalities, provide an initial translational link between progerin-targeted intervention and measurable structural changes in human skin.

Although the single-arm design and multi-ingredient formulation do not allow the specific contribution of Progerinin to be determined, the substantial dermal density response provides a strong rationale for further controlled evaluation of Progerinin as a potential contributor to the observed effects. Randomized studies comparing of otherwise identical formulations with and without Progerinin, ideally combined with direct assessment of cutaneous progerin and extracellular matrix markers, will be important for clarifying its specific contribution and biological mechanism.

## Data Availability

All data produced in the present study are available upon reasonable request to the authors

## Acknowledgment

This research was supported by Korea Drug Development Fund funded by Ministry of Science and ICT, Ministry of Trade, Industry, and Energy, and Ministry of Health and Welfare (RS-2023-00258708).

## References

1. McClintock D, Ratner D, Lokuge M, Owens DM, Gordon LB, Collins FS, and Djabali K. The mutant form of lamin A that causes Hutchinson-Gilford progeria is a biomarker of cellular aging in human skin. 2007;2(12):e1269.

2. Kang S, Park S, Woo T-G, Kim B-H, and Park B-J. Progerin expression in humans: implications for natural ageing. 2026;112164.

3. Draelos ZD, Diaz I, Namkoong J, Wu J, and Boyd T. Efficacy evaluation of a topical hyaluronic acid serum in facial photoaging. 2021;11(4):1385–94.

4. Cole MA, Quan T, Voorhees JJ, and Fisher GJ. Extracellular matrix regulation of fibroblast function: redefining our perspective on skin aging: Cole MA et al. 2018;12(1):35–43.

5. Rovero P, Malgapo DMH, Sparavigna A, Beilin G, Wong V, and Lao MP. The Clinical Evidence-Based Paradigm of Topical Anti-Aging Skincare Formulations Enriched with Bio-Active Peptide SA1-III (KP1) as Collagen Modulator: From Bench to Bedside [Corrigendum]. 2023;16:1855–6.

6. Alves PLM, Nieri V, Moreli F de C, Constantino E, de Souza J, Oshima-Franco Y, and Grotto D. Unveiling new horizons: advancing technologies in cosmeceuticals for anti-aging solutions. 2024;29(20):4890.

7. Wlaschek M, Maity P, Makrantonaki E, and Scharffetter-Kochanek K. Connective tissue and fibroblast senescence in skin aging. 2021;141(4):985–92.

8. Brusini R, Peslerbe L, Vantou C, Meriaux P, Clark Loeser LR, and Faivre J. Crosslinked HA in Topical Application: The Biological Benefits of RHA Serum on Aging Signs. 2026;25(7):e71065.

9. Lee S-J, Jung Y-S, Yoon M-H, Kang S, Oh A-Y, Lee J-H, Jun S-Y, Woo T-G, Chun H-Y, and Kim SK. Interruption of progerin–lamin A/C binding ameliorates Hutchinson-Gilford progeria syndrome phenotype. 2016;126(10):3879–93.

10. Madaan P, Sikka P, and Malik DS. Cosmeceutical aptitudes of niacinamide: a review. 2021;16(3):196–208.

11. Shin J-W, Kwon S-H, Choi J-Y, Na J-I, Huh C-H, Choi H-R, and Park K-C. Molecular mechanisms of dermal aging and antiaging approaches. 2019;20(9):2126.

12. Qi M, Pitta P, Wegner K, Kristof B, Feng Y, Raddatz G, Rodríguez-Paredes M, Gallinger J, Wanitphakdeedecha R, and Winnefeld M. Epigenetic Skin Aging and Its Reversal to Improve Skin Longevity across Ethnicities and Phototypes Using a Dihydromyricetin-Containing Serum: Results from a Prospective, Single-Cohort Study. 2026;1–16.

13. Gniadecka, M. Effects of ageing on dermal echogenicity. Skin Research and Technology 7.3 (2001) 204–207.

14. Zhang J, Yu H, Man M, and Hu L. Aging in the dermis: Fibroblast senescence and its significance. 2024;23(2):e14054.

15. Rawlings AV and Harding CR. Moisturization and skin barrier function. 2004;17:43–8.

16. Kang S, Yoon M-H, Ahn J, Kim J-E, Kim SY, Kang SY, Joo J, Park S, Cho J-H, and Woo T-G. Progerinin, an optimized progerin-lamin A binding inhibitor, ameliorates premature senescence phenotypes of Hutchinson-Gilford progeria syndrome. 2021;4(1):5.

17. Gruber F, Kremslehner C, Eckhart L, and Tschachler E. Cell aging and cellular senescence in skin aging—Recent advances in fibroblast and keratinocyte biology. 2020;130:110780.

18. Baumann L. Skin ageing and its treatment. 2007;211(2):241–51.

19. Potekaev NN, Borzykh OB, Medvedev GV, Pushkin DV, Petrova MM, Petrov AV, Dmitrenko DV, Karpova EI, Demina OM, and Shnayder NA. The role of extracellular matrix in skin wound healing. 2021;10(24):5947.

20. Papaccio F, D′ Arino A, Caputo S, and Bellei B. Focus on the contribution of oxidative stress in skin aging. 2022;11(6):1121.

21. Fisher GJ and Voorhees JJ. Molecular mechanisms of photoaging and its prevention by retinoic acid: ultraviolet irradiation induces MAP kinase signal transduction cascades that induce Ap-1-regulated matrix metalloproteinases that degrade human skin in vivo. Journal of Investigative Dermatology Symposium Proceedings, Vol 3. Elsevier; 1998.

22. Scaffidi P and Misteli T. Lamin A-dependent nuclear defects in human aging. 2006;312(5776):1059–63.

23. Kang S, Yoon M-H, Lee S-J, Ahn J, Yi SA, Nam KH, Park S, Woo T-G, Cho J-H, and Lee J. Human WRN is an intrinsic inhibitor of progerin, abnormal splicing product of lamin A. 2021;11(1):9122.

24. Orioli D and Dellambra E. Epigenetic regulation of skin cells in natural aging and premature aging diseases. 2018;7(12):268.

25. Naidoo K and Birch-Machin MA. Oxidative stress and ageing: the influence of environmental pollution, sunlight and diet on skin. 2017;4(1):4.

26. Eriksson M, Brown WT, Gordon LB, Glynn MW, Singer J, Scott L, Erdos MR, Robbins CM, Moses TY, and Berglund P. Recurrent de novo point mutations in lamin A cause Hutchinson–Gilford progeria syndrome. 2003;423(6937):293–8.

27. Bravo, Bruna, et al. Benefits of topical hyaluronic acid for skin quality and signs of skin aging From literature review to clinical evidence. Dermatologic therapy 35.12 (2022) e15903.

28. Fisher GJ, Talwar HS, Lin J, and Voorhees JJ. Molecular mechanisms of photoaging in human skin in vivo and their prevention by all trans retinoic acid. 1999;69(2):154–7.

29. Tobin DJ. Introduction to skin aging. 2017;26(1):37–46.

30. Kim KH, Kim Y-S, Lee S, and An S. The effect of three-dimensional cultured adipose tissue-derived mesenchymal stem cell–conditioned medium and the antiaging effect of cosmetic products containing the medium. 2019;4(1):1.

31. Krutmann J, Bouloc A, Sore G, Bernard BA, and Passeron T. The skin aging exposome. 2017;85(3):152–61.

32. Nam G, Lee HW, Jang J, Kim CH, and Kim K. Novel conformation of hyaluronic acid with improved cosmetic efficacy. 2023;22(4):1312–20.

33. Farage MA, Miller KW, Elsner P, and Maibach HI. Intrinsic and extrinsic factors in skin ageing: a review. 2008;30(2):87–95.

34. Bednarski IA, Ciążyńska M, Kabziński J, Majsterek I, Sobolewska-Sztychny D, Narbutt J, and Lesiak A. More Than Skin Deep–the Effects of Ultraviolet Radiation on Cathepsin K and Progerin Expression in Cultured Dermal Fibroblasts. 2021;1561–8.

35. Takeuchi H and Rünger TM. Longwave UV light induces the aging-associated progerin. 2013;133(7):1857–62.

36. Kameyama K, Sakai C, Kondoh S, Yonemoto K, Nishiyama S, Tagawa M, Murata T, Ohnuma T, Quigley J, and Dorsky A. Inhibitory effect of magnesium L-ascorbyl-2-phosphate (VC-PMG) on melanogenesis in vitro and in vivo. 1996;34(1):29–33.

37. Bissett DL, Oblong JE, and Berge CA. Niacinamide: AB vitamin that improves aging facial skin appearance. 2005;31:860–6.

38. Kim B-H, Chung Y-H, Woo T-G, Kang S-M, Park S, and Park B-J. Progerin, an aberrant spliced form of lamin A, is a potential therapeutic target for HGPS. 2023;12(18):2299.

39. Kang S, Seo S, Song EJ, Kweon O, Jo A, Park S, Woo T-G, Kim B-H, Oh GT, and Park B-J. Progerinin, an inhibitor of Progerin, alleviates cardiac abnormalities in a model mouse of Hutchinson– Gilford progeria syndrome. 2023;12(9):1232.

40. He X, Gao X, Guo Y, and Xie W. Research progress on bioactive factors against skin aging. 2024;25(7):3797.

