## Supplementary Figure 1 for "Short-Term Changes in Dermal Density Following Topical Application of a Progerinin-Containing Serum: An Exploratory Clinical Study"

### Slide 1
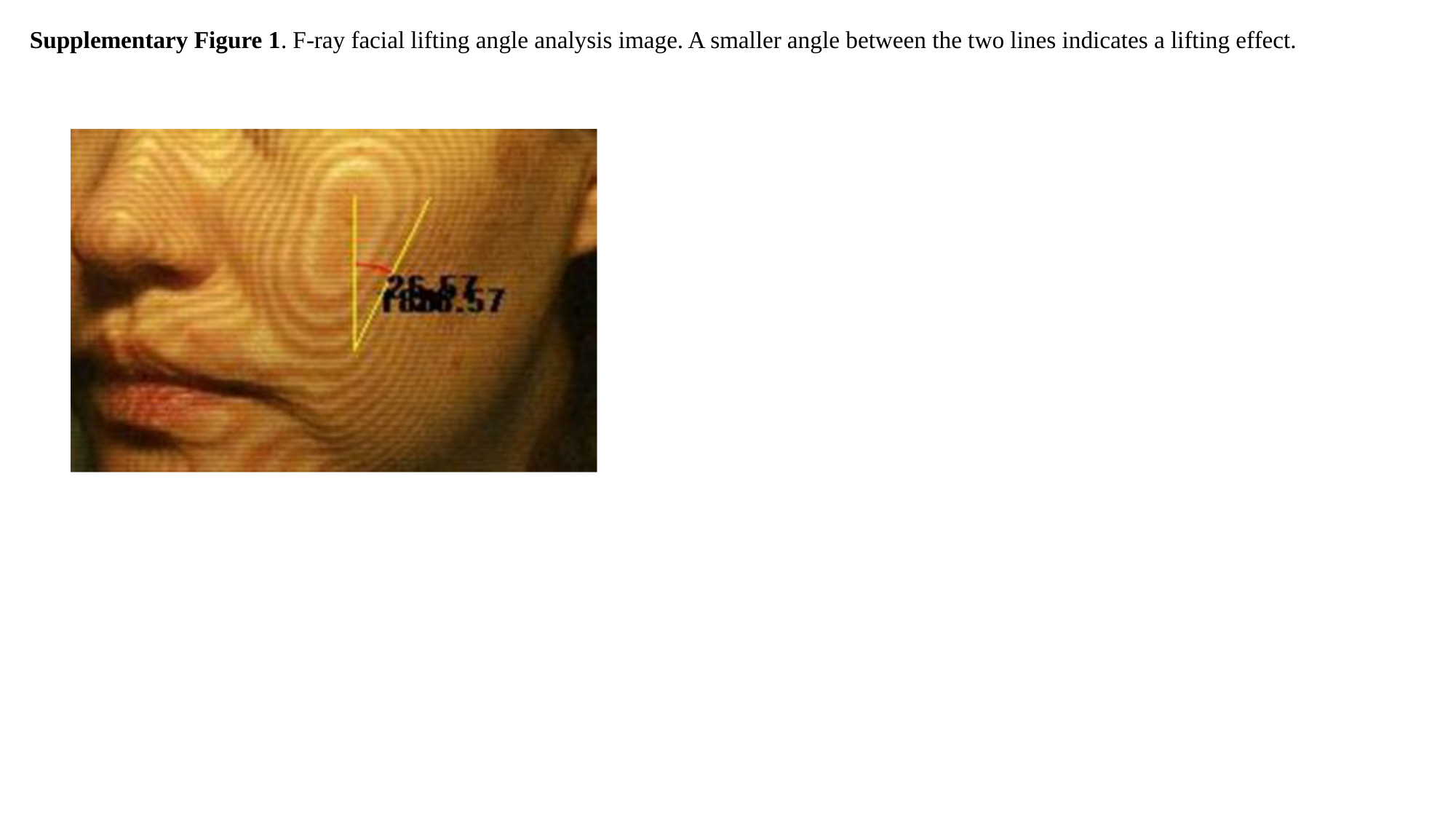

Supplementary Figure 1. F-ray facial lifting angle analysis image. A smaller angle between the two lines indicates a lifting effect.
