## Supplementary material for "Short-Term Changes in Dermal Density Following Topical Application of a Progerinin-Containing Serum: An Exploratory Clinical Study": Tables

### Slide 1
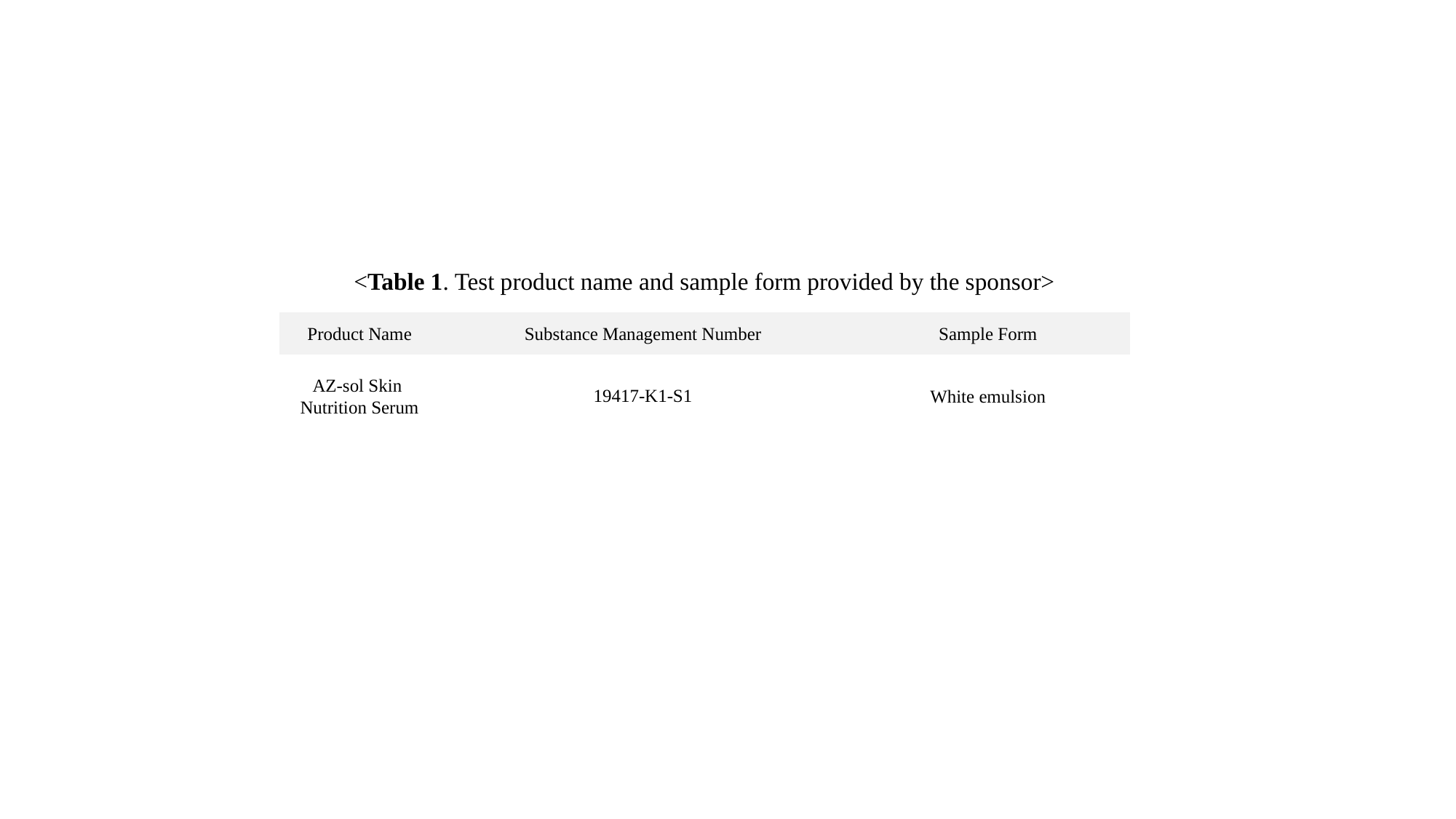

<Table 1. Test product name and sample form provided by the sponsor>
| Product Name | Substance Management Number | Sample Form |
| --- | --- | --- |
| AZ-sol Skin Nutrition Serum | 19417-K1-S1 | White emulsion |

### Slide 2
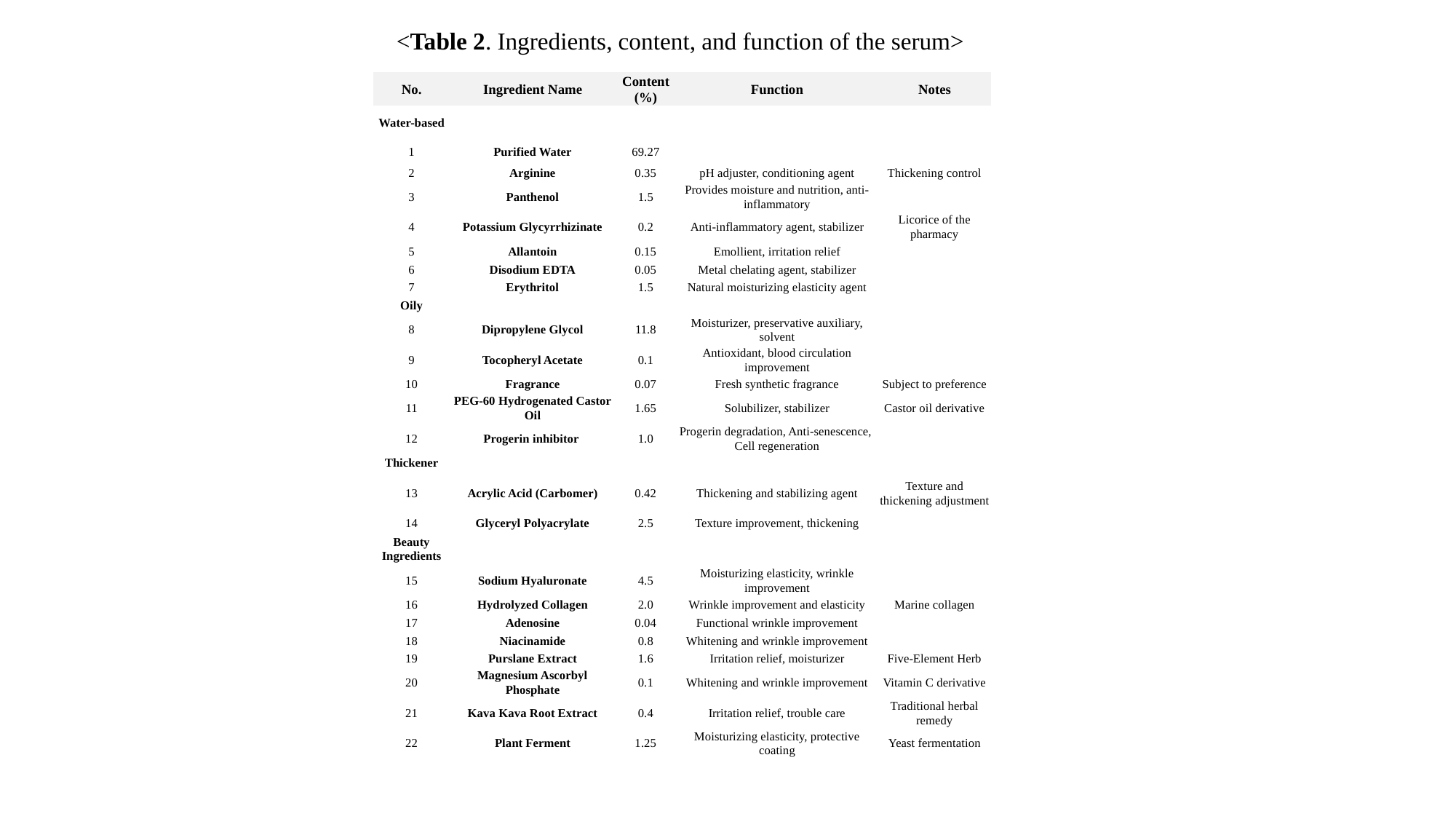

<Table 2. Ingredients, content, and function of the serum>
| No. | Ingredient Name | Content (%) | Function | Notes |
| --- | --- | --- | --- | --- |
| Water-based | | | | |
| 1 | Purified Water | 69.27 | | |
| 2 | Arginine | 0.35 | pH adjuster, conditioning agent | Thickening control |
| 3 | Panthenol | 1.5 | Provides moisture and nutrition, anti-inflammatory | |
| 4 | Potassium Glycyrrhizinate | 0.2 | Anti-inflammatory agent, stabilizer | Licorice of the pharmacy |
| 5 | Allantoin | 0.15 | Emollient, irritation relief | |
| 6 | Disodium EDTA | 0.05 | Metal chelating agent, stabilizer | |
| 7 | Erythritol | 1.5 | Natural moisturizing elasticity agent | |
| Oily | | | | |
| 8 | Dipropylene Glycol | 11.8 | Moisturizer, preservative auxiliary, solvent | |
| 9 | Tocopheryl Acetate | 0.1 | Antioxidant, blood circulation improvement | |
| 10 | Fragrance | 0.07 | Fresh synthetic fragrance | Subject to preference |
| 11 | PEG-60 Hydrogenated Castor Oil | 1.65 | Solubilizer, stabilizer | Castor oil derivative |
| 12 | Progerin inhibitor | 1.0 | Progerin degradation, Anti-senescence, Cell regeneration | |
| Thickener | | | | |
| 13 | Acrylic Acid (Carbomer) | 0.42 | Thickening and stabilizing agent | Texture and thickening adjustment |
| 14 | Glyceryl Polyacrylate | 2.5 | Texture improvement, thickening | |
| Beauty Ingredients | | | | |
| 15 | Sodium Hyaluronate | 4.5 | Moisturizing elasticity, wrinkle improvement | |
| 16 | Hydrolyzed Collagen | 2.0 | Wrinkle improvement and elasticity | Marine collagen |
| 17 | Adenosine | 0.04 | Functional wrinkle improvement | |
| 18 | Niacinamide | 0.8 | Whitening and wrinkle improvement | |
| 19 | Purslane Extract | 1.6 | Irritation relief, moisturizer | Five-Element Herb |
| 20 | Magnesium Ascorbyl Phosphate | 0.1 | Whitening and wrinkle improvement | Vitamin C derivative |
| 21 | Kava Kava Root Extract | 0.4 | Irritation relief, trouble care | Traditional herbal remedy |
| 22 | Plant Ferment | 1.25 | Moisturizing elasticity, protective coating | Yeast fermentation |

### Slide 3
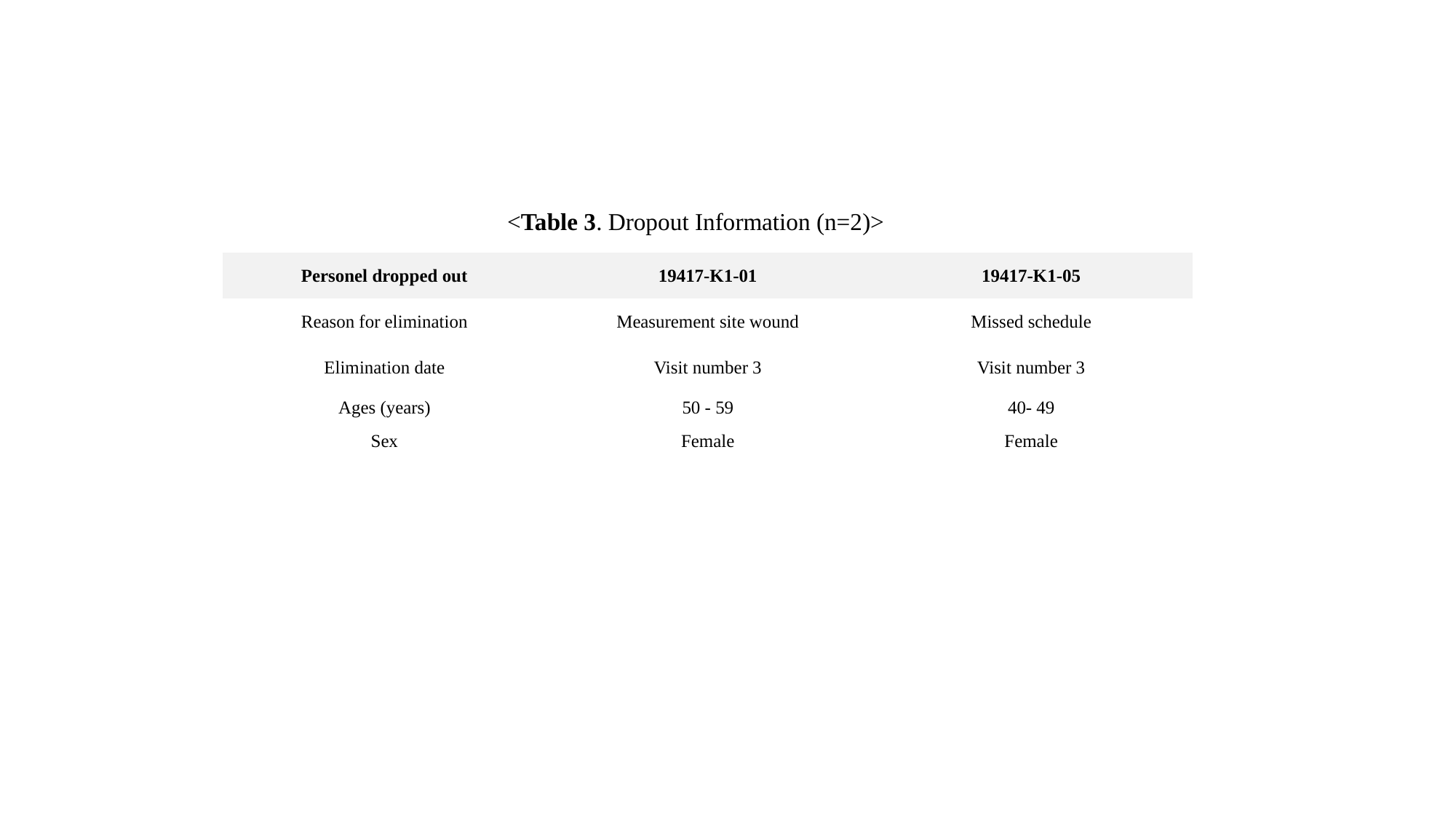

<Table 3. Dropout Information (n=2)>
| Personel dropped out | 19417-K1-01 | 19417-K1-05 |
| --- | --- | --- |
| Reason for elimination | Measurement site wound | Missed schedule |
| Elimination date | Visit number 3 | Visit number 3 |
| Ages (years) | 50 - 59 | 40- 49 |
| Sex | Female | Female |

### Slide 4
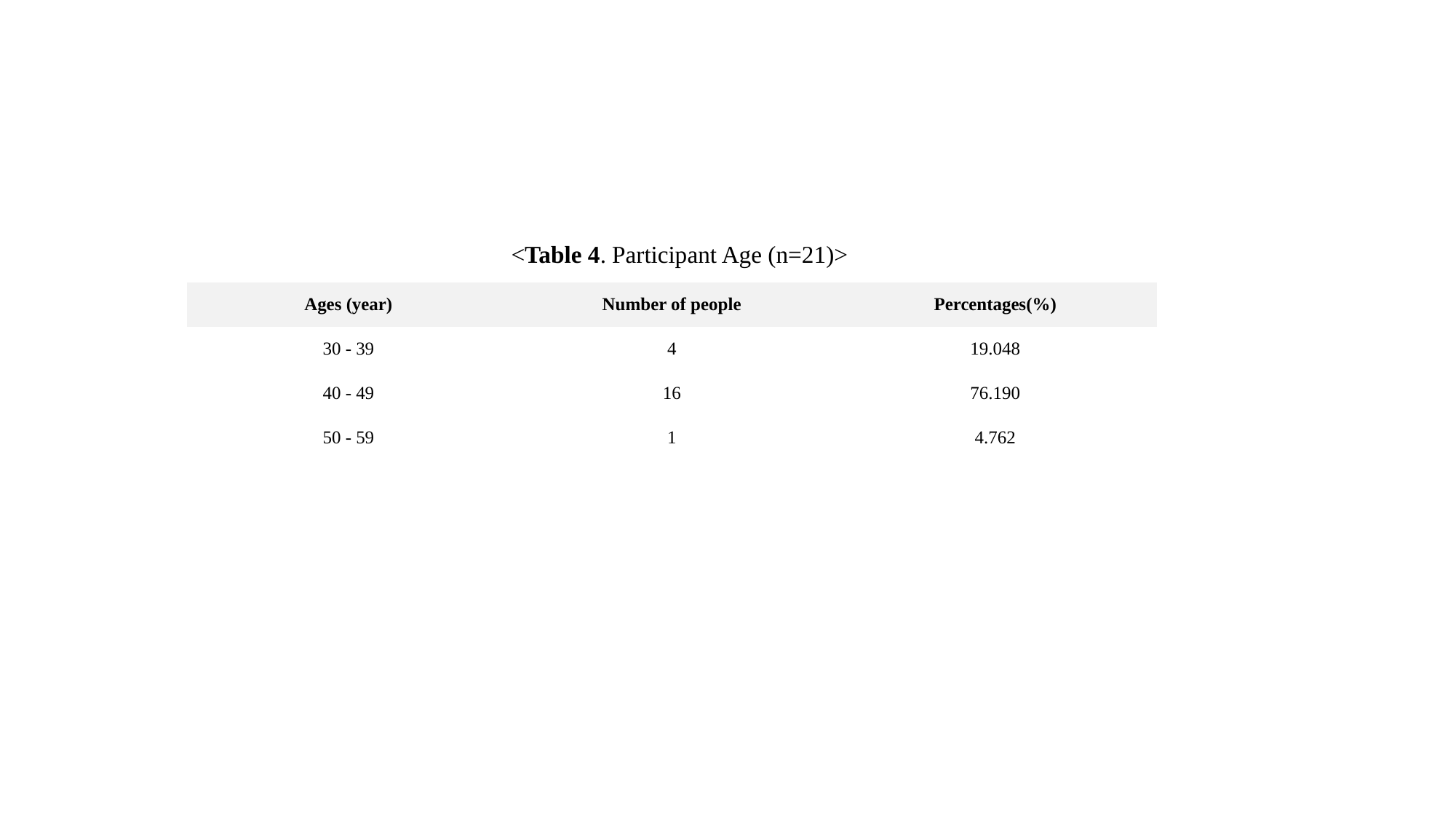

<Table 4. Participant Age (n=21)>
| Ages (year) | Number of people | Percentages(%) |
| --- | --- | --- |
| 30 - 39 | 4 | 19.048 |
| 40 - 49 | 16 | 76.190 |
| 50 - 59 | 1 | 4.762 |

### Slide 5
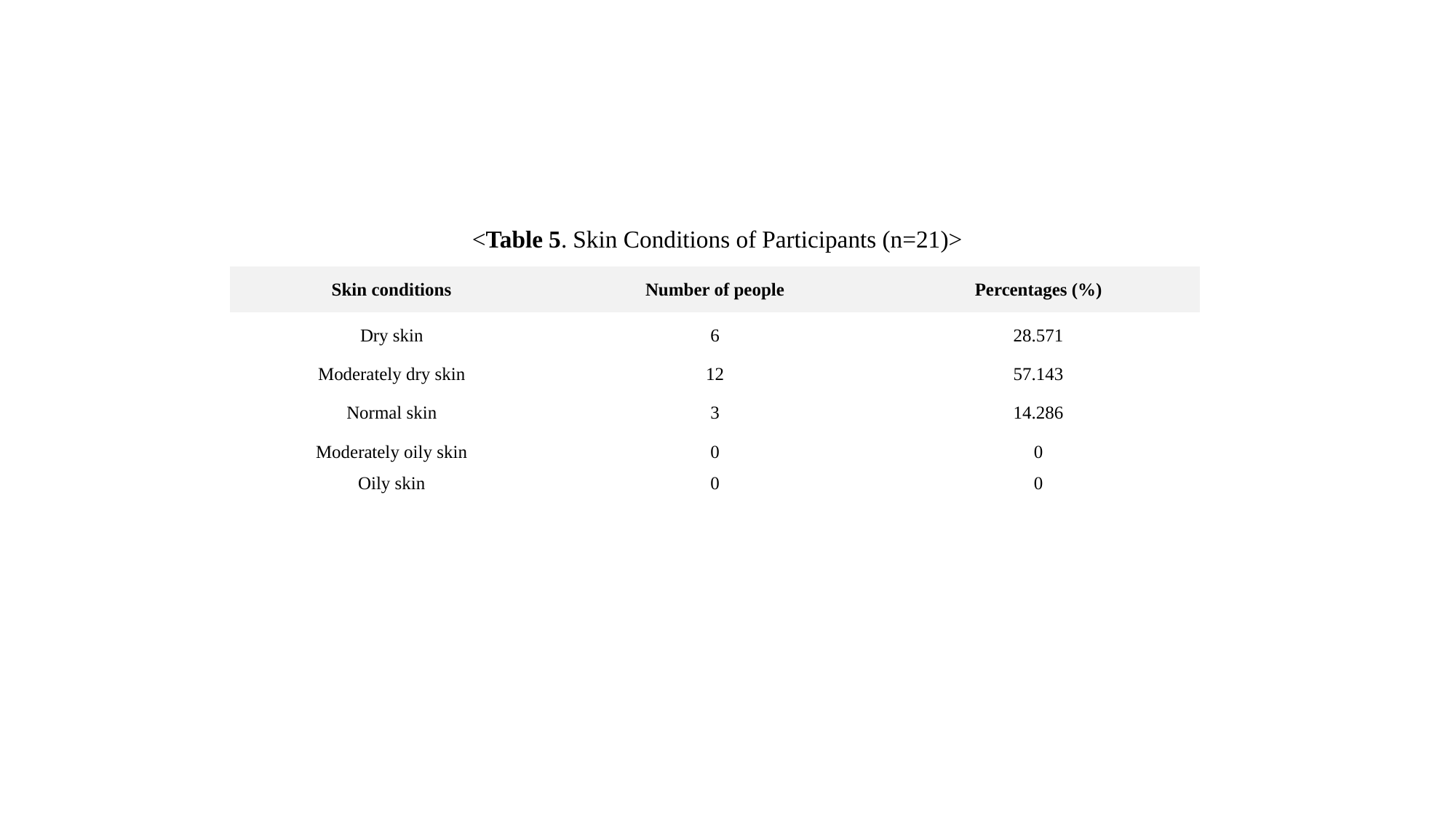

<Table 5. Skin Conditions of Participants (n=21)>
| Skin conditions | Number of people | Percentages (%) |
| --- | --- | --- |
| Dry skin | 6 | 28.571 |
| Moderately dry skin | 12 | 57.143 |
| Normal skin | 3 | 14.286 |
| Moderately oily skin | 0 | 0 |
| Oily skin | 0 | 0 |

### Slide 6
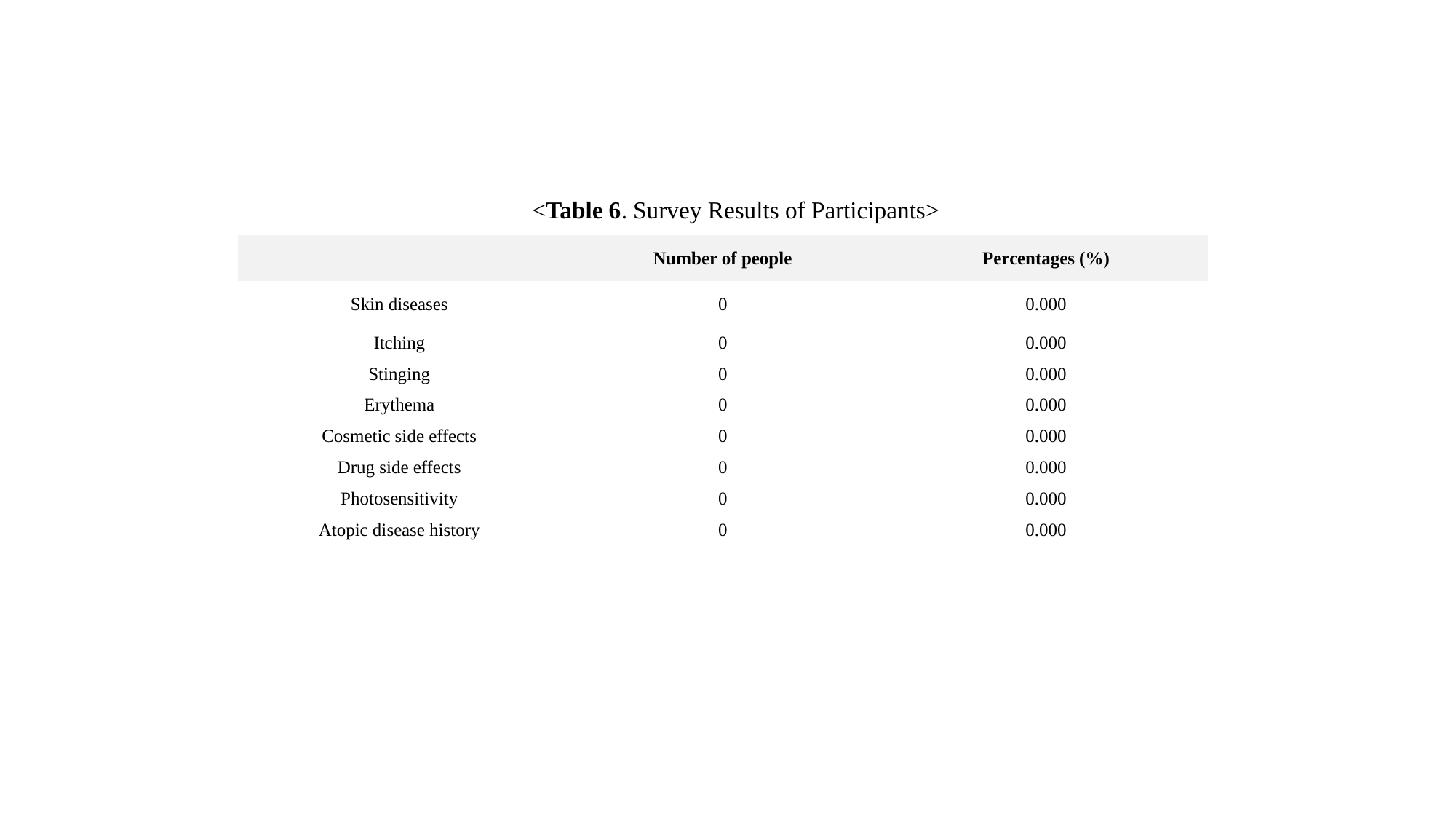

<Table 6. Survey Results of Participants>
| | Number of people | Percentages (%) |
| --- | --- | --- |
| Skin diseases | 0 | 0.000 |
| Itching | 0 | 0.000 |
| Stinging | 0 | 0.000 |
| Erythema | 0 | 0.000 |
| Cosmetic side effects | 0 | 0.000 |
| Drug side effects | 0 | 0.000 |
| Photosensitivity | 0 | 0.000 |
| Atopic disease history | 0 | 0.000 |

### Slide 7
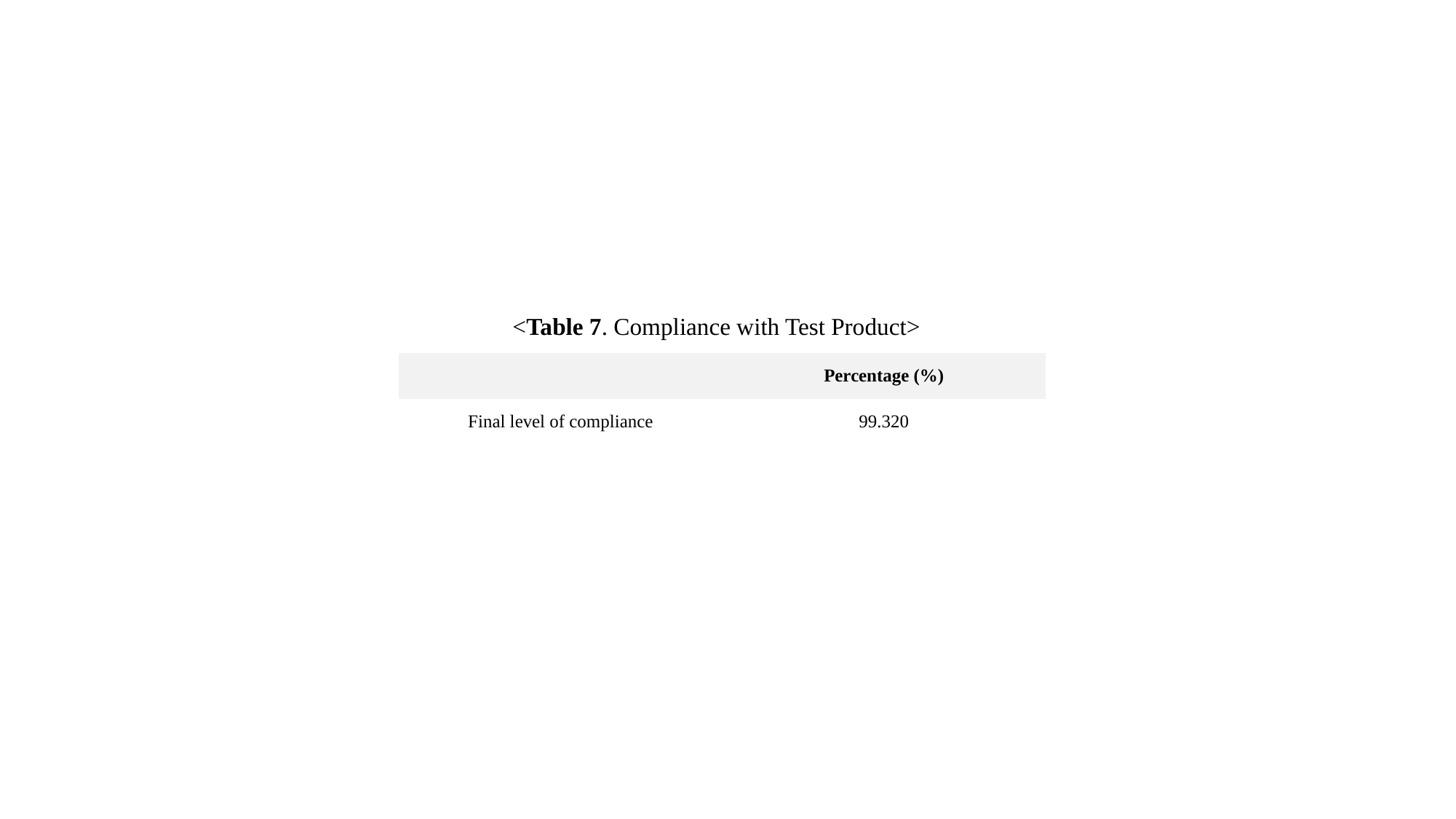

<Table 7. Compliance with Test Product>
| | Percentage (%) |
| --- | --- |
| Final level of compliance | 99.320 |

### Slide 8
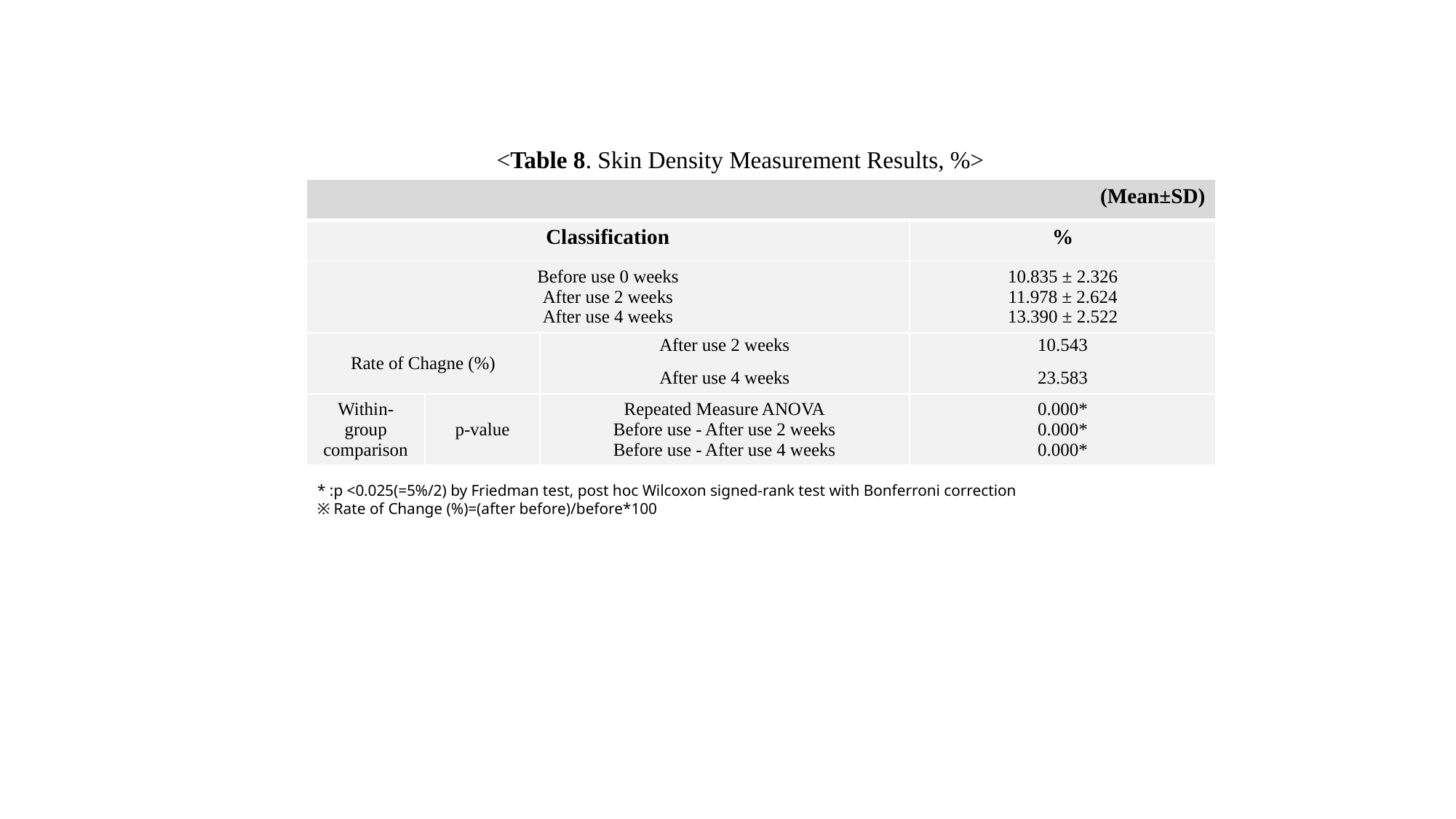

<Table 8. Skin Density Measurement Results, %>
| (Mean±SD) | | | |
| --- | --- | --- | --- |
| Classification | | | % |
| Before use 0 weeks After use 2 weeks After use 4 weeks | | | 10.835 ± 2.326 11.978 ± 2.624 13.390 ± 2.522 |
| Rate of Chagne (%) | | After use 2 weeks After use 4 weeks | 10.543 23.583 |
| Within-group comparison | p-value | Repeated Measure ANOVA Before use - After use 2 weeks Before use - After use 4 weeks | 0.000\* 0.000\* 0.000\* |
* :p <0.025(=5%/2) by Friedman test, post hoc Wilcoxon signed-rank test with Bonferroni correction
※ Rate of Change (%)=(after before)/before*100

### Slide 9
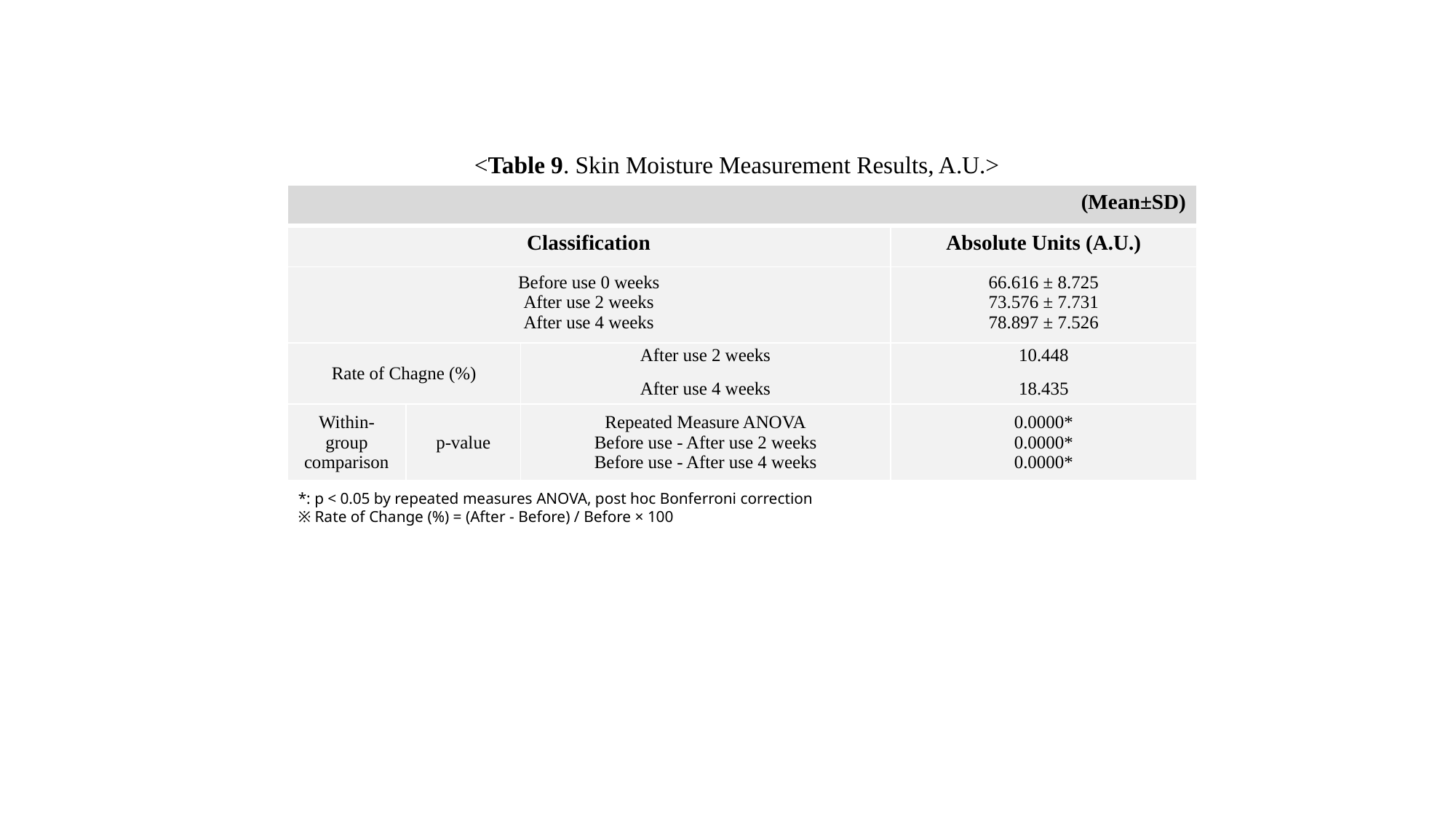

<Table 9. Skin Moisture Measurement Results, A.U.>
| (Mean±SD) | | | |
| --- | --- | --- | --- |
| Classification | | | Absolute Units (A.U.) |
| Before use 0 weeks After use 2 weeks After use 4 weeks | | | 66.616 ± 8.725 73.576 ± 7.731 78.897 ± 7.526 |
| Rate of Chagne (%) | | After use 2 weeks After use 4 weeks | 10.448 18.435 |
| Within-group comparison | p-value | Repeated Measure ANOVA Before use - After use 2 weeks Before use - After use 4 weeks | 0.0000\* 0.0000\* 0.0000\* |
*: p < 0.05 by repeated measures ANOVA, post hoc Bonferroni correction
※ Rate of Change (%) = (After - Before) / Before × 100

### Slide 10
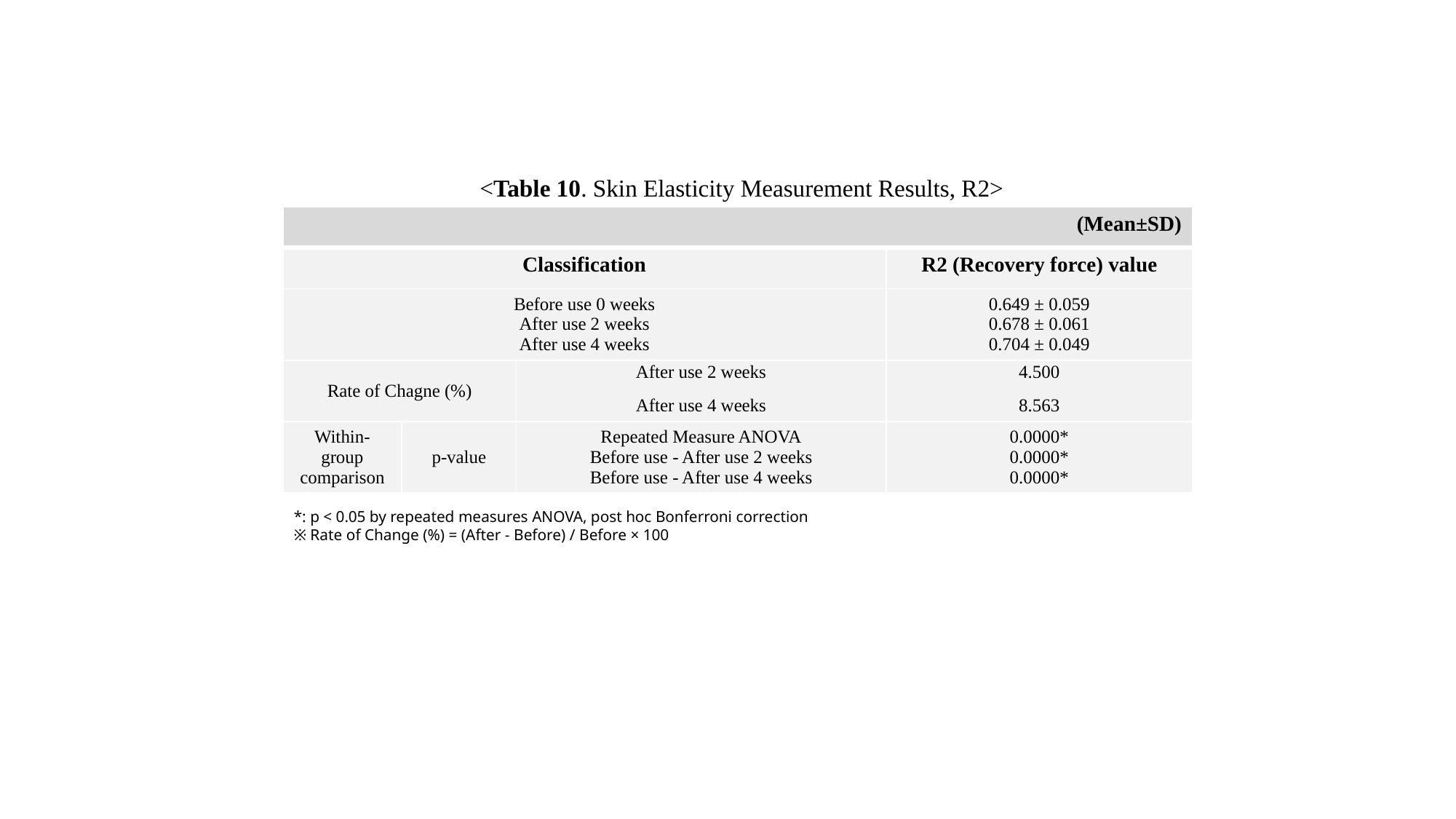

<Table 10. Skin Elasticity Measurement Results, R2>
| (Mean±SD) | | | |
| --- | --- | --- | --- |
| Classification | | | R2 (Recovery force) value |
| Before use 0 weeks After use 2 weeks After use 4 weeks | | | 0.649 ± 0.059 0.678 ± 0.061 0.704 ± 0.049 |
| Rate of Chagne (%) | | After use 2 weeks After use 4 weeks | 4.500 8.563 |
| Within-group comparison | p-value | Repeated Measure ANOVA Before use - After use 2 weeks Before use - After use 4 weeks | 0.0000\* 0.0000\* 0.0000\* |
*: p < 0.05 by repeated measures ANOVA, post hoc Bonferroni correction
※ Rate of Change (%) = (After - Before) / Before × 100

### Slide 11
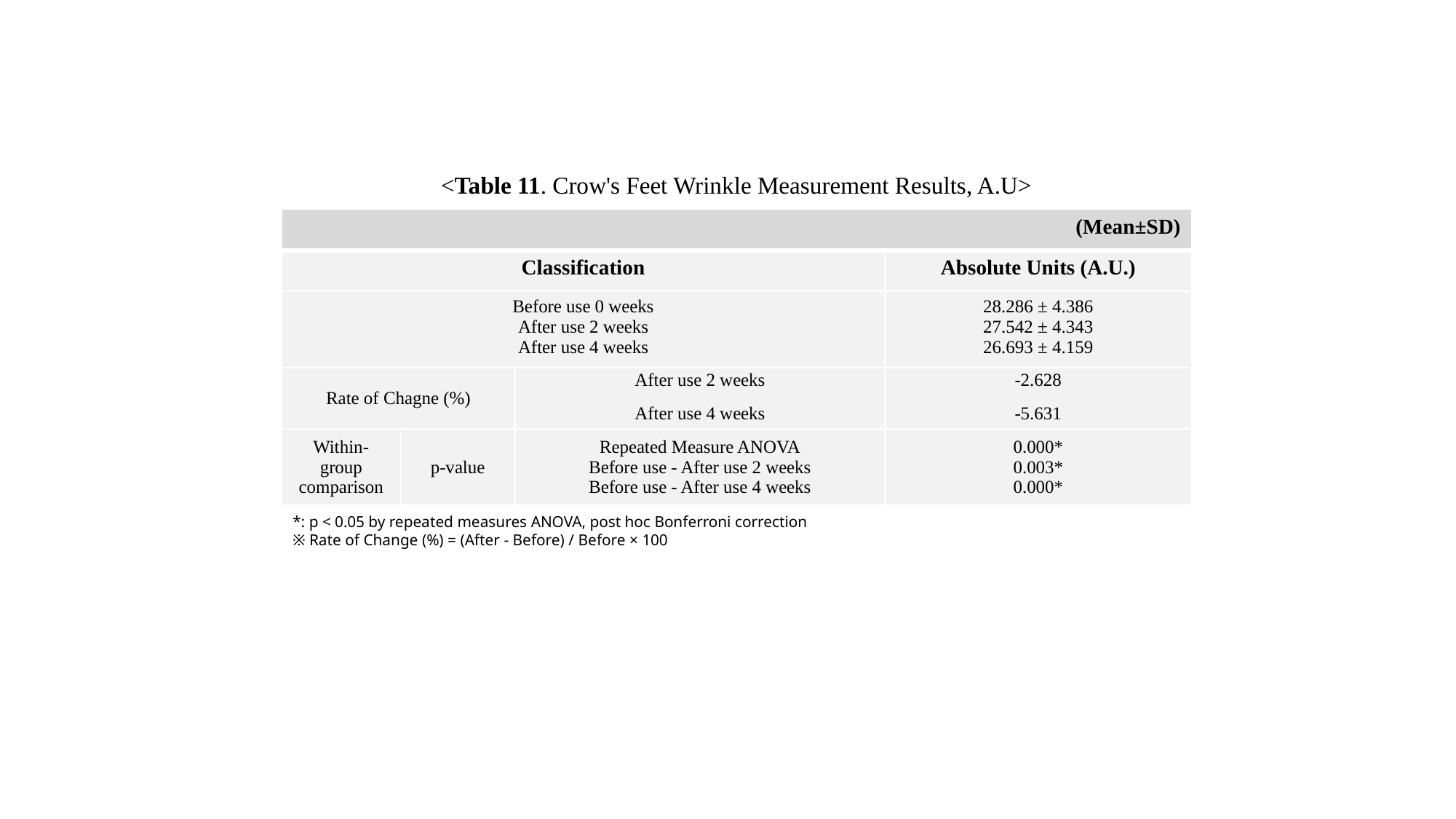

<Table 11. Crow's Feet Wrinkle Measurement Results, A.U>
| (Mean±SD) | | | |
| --- | --- | --- | --- |
| Classification | | | Absolute Units (A.U.) |
| Before use 0 weeks After use 2 weeks After use 4 weeks | | | 28.286 ± 4.386 27.542 ± 4.343 26.693 ± 4.159 |
| Rate of Chagne (%) | | After use 2 weeks After use 4 weeks | -2.628 -5.631 |
| Within-group comparison | p-value | Repeated Measure ANOVA Before use - After use 2 weeks Before use - After use 4 weeks | 0.000\* 0.003\* 0.000\* |
*: p < 0.05 by repeated measures ANOVA, post hoc Bonferroni correction
※ Rate of Change (%) = (After - Before) / Before × 100

### Slide 12
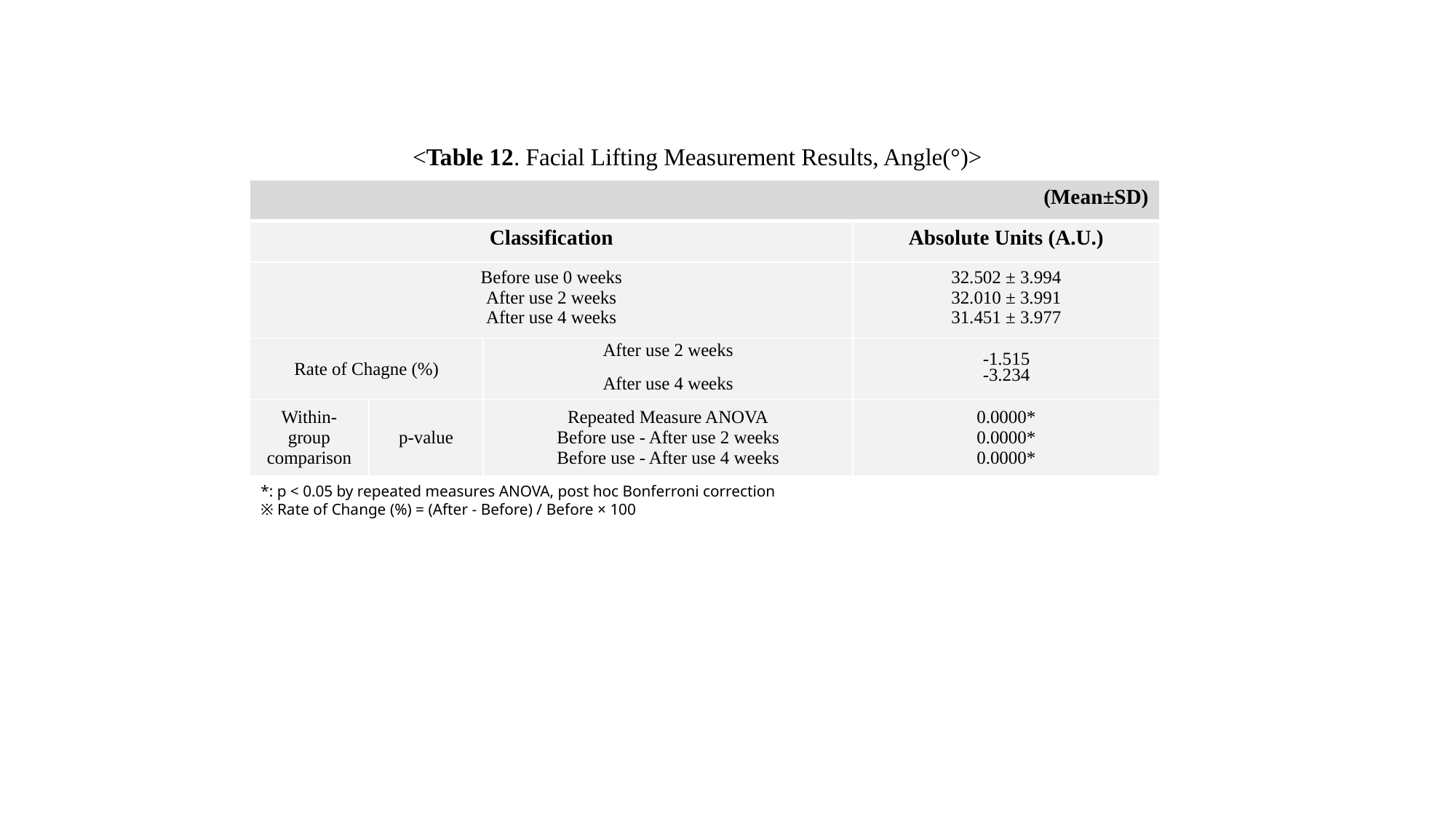

<Table 12. Facial Lifting Measurement Results, Angle(°)>
| (Mean±SD) | | | |
| --- | --- | --- | --- |
| Classification | | | Absolute Units (A.U.) |
| Before use 0 weeks After use 2 weeks After use 4 weeks | | | 32.502 ± 3.994 32.010 ± 3.991 31.451 ± 3.977 |
| Rate of Chagne (%) | | After use 2 weeks After use 4 weeks | -1.515 -3.234 |
| Within-group comparison | p-value | Repeated Measure ANOVA Before use - After use 2 weeks Before use - After use 4 weeks | 0.0000\* 0.0000\* 0.0000\* |
*: p < 0.05 by repeated measures ANOVA, post hoc Bonferroni correction
※ Rate of Change (%) = (After - Before) / Before × 100

### Slide 13
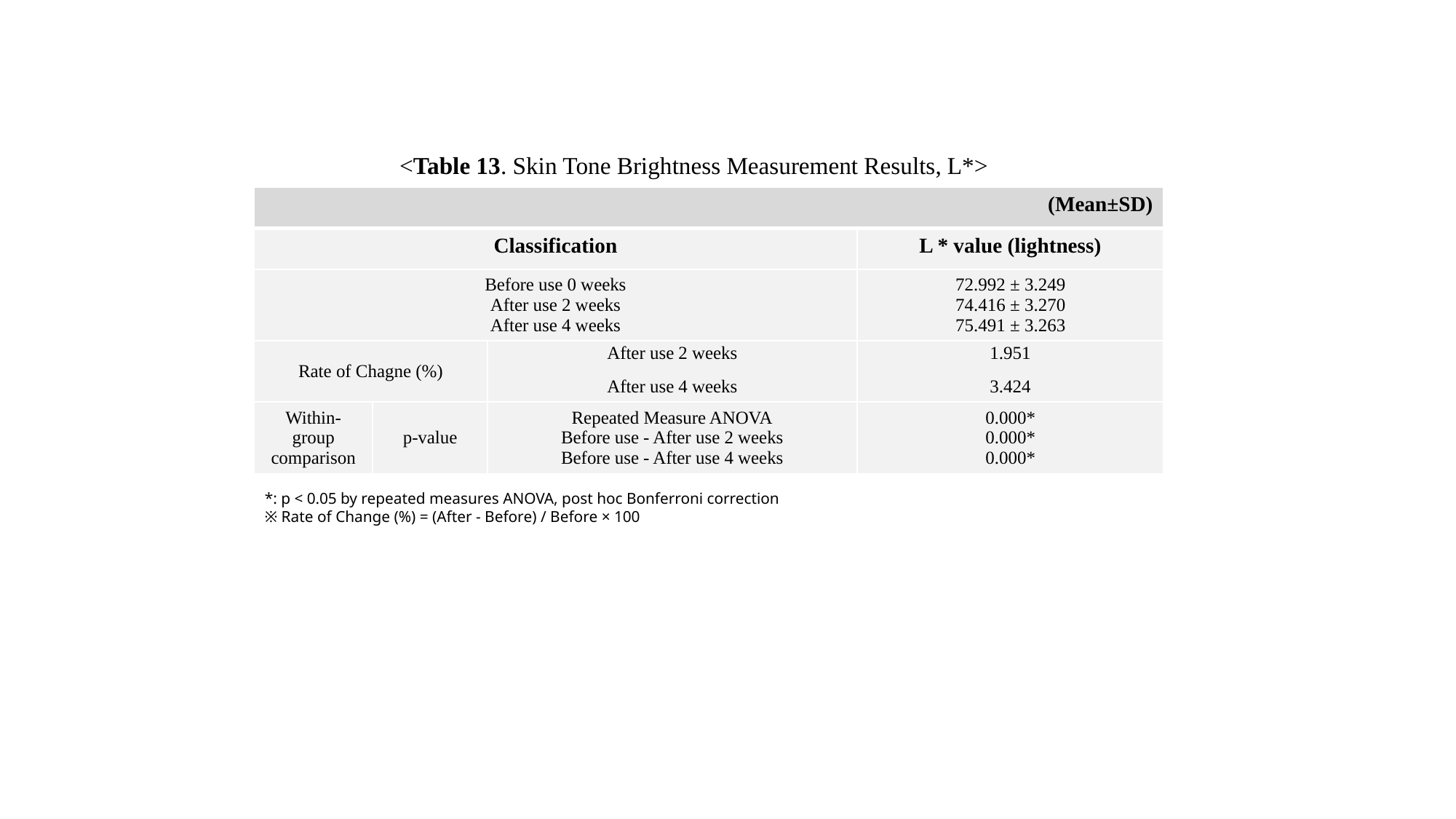

<Table 13. Skin Tone Brightness Measurement Results, L*>
| (Mean±SD) | | | |
| --- | --- | --- | --- |
| Classification | | | L \* value (lightness) |
| Before use 0 weeks After use 2 weeks After use 4 weeks | | | 72.992 ± 3.249 74.416 ± 3.270 75.491 ± 3.263 |
| Rate of Chagne (%) | | After use 2 weeks After use 4 weeks | 1.951 3.424 |
| Within-group comparison | p-value | Repeated Measure ANOVA Before use - After use 2 weeks Before use - After use 4 weeks | 0.000\* 0.000\* 0.000\* |
*: p < 0.05 by repeated measures ANOVA, post hoc Bonferroni correction
※ Rate of Change (%) = (After - Before) / Before × 100

### Slide 14
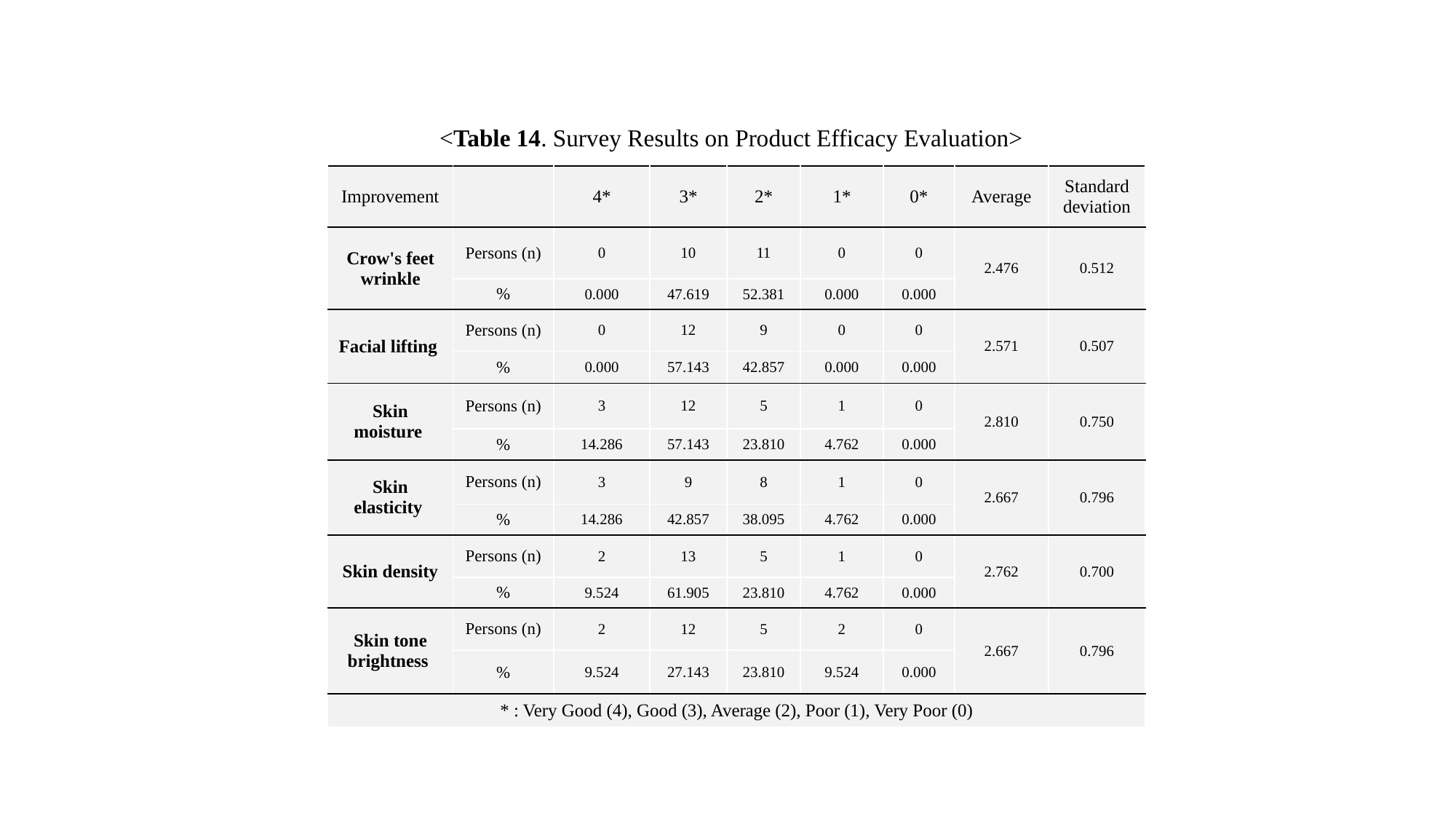

<Table 14. Survey Results on Product Efficacy Evaluation>
| Improvement | | 4\* | 3\* | 2\* | 1\* | 0\* | Average | Standard deviation |
| --- | --- | --- | --- | --- | --- | --- | --- | --- |
| Crow's feet wrinkle | Persons (n) | 0 | 10 | 11 | 0 | 0 | 2.476 | 0.512 |
| | % | 0.000 | 47.619 | 52.381 | 0.000 | 0.000 | | |
| Facial lifting | Persons (n) | 0 | 12 | 9 | 0 | 0 | 2.571 | 0.507 |
| | % | 0.000 | 57.143 | 42.857 | 0.000 | 0.000 | | |
| Skin moisture | Persons (n) | 3 | 12 | 5 | 1 | 0 | 2.810 | 0.750 |
| | % | 14.286 | 57.143 | 23.810 | 4.762 | 0.000 | | |
| Skin elasticity | Persons (n) | 3 | 9 | 8 | 1 | 0 | 2.667 | 0.796 |
| | % | 14.286 | 42.857 | 38.095 | 4.762 | 0.000 | | |
| Skin density | Persons (n) | 2 | 13 | 5 | 1 | 0 | 2.762 | 0.700 |
| | % | 9.524 | 61.905 | 23.810 | 4.762 | 0.000 | | |
| Skin tone brightness | Persons (n) | 2 | 12 | 5 | 2 | 0 | 2.667 | 0.796 |
| | % | 9.524 | 27.143 | 23.810 | 9.524 | 0.000 | | |
| \* : Very Good (4), Good (3), Average (2), Poor (1), Very Poor (0) | | | | | | | | |

### Slide 15
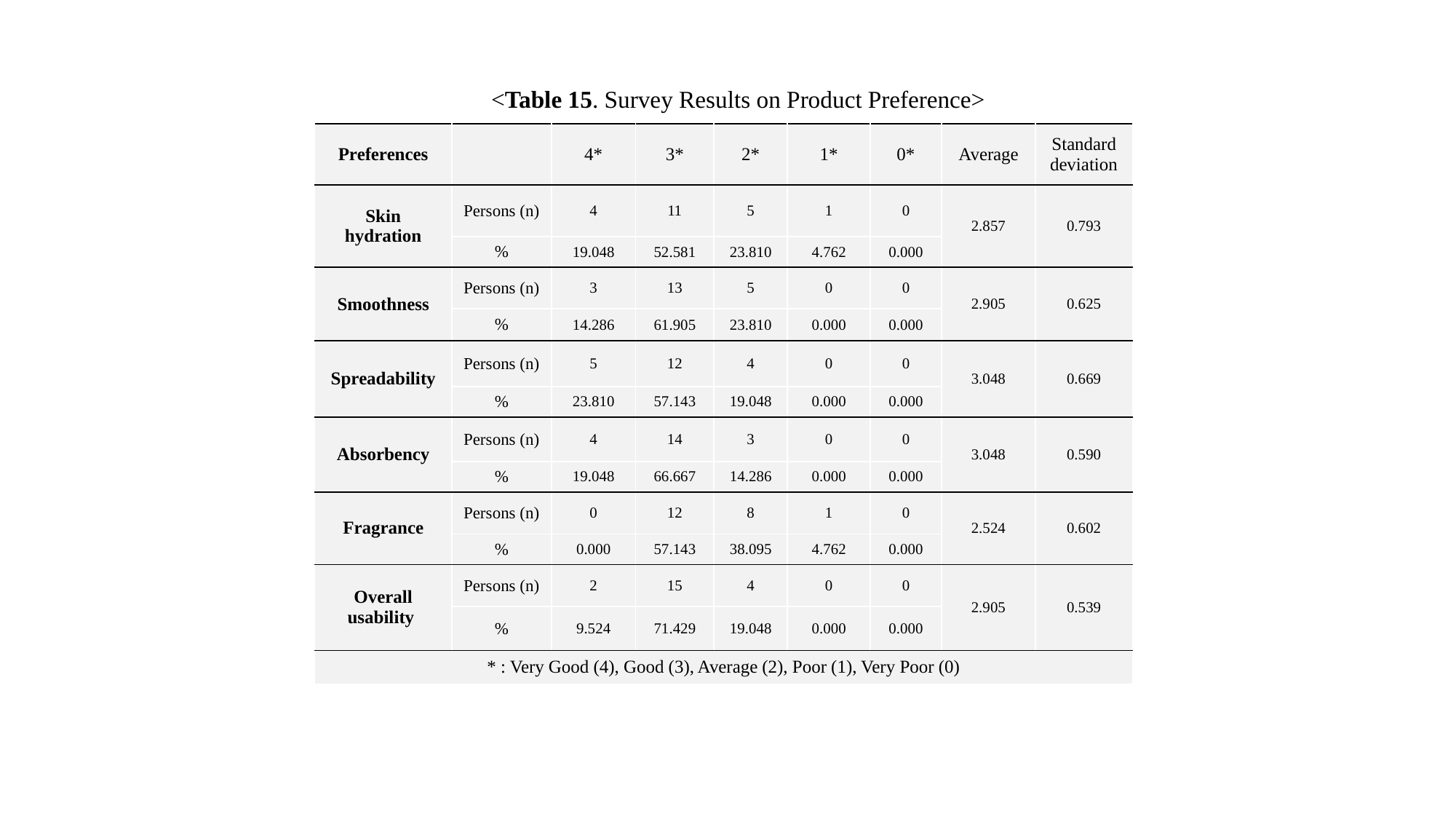

<Table 15. Survey Results on Product Preference>
| Preferences | | 4\* | 3\* | 2\* | 1\* | 0\* | Average | Standard deviation |
| --- | --- | --- | --- | --- | --- | --- | --- | --- |
| Skin hydration | Persons (n) | 4 | 11 | 5 | 1 | 0 | 2.857 | 0.793 |
| | % | 19.048 | 52.581 | 23.810 | 4.762 | 0.000 | | |
| Smoothness | Persons (n) | 3 | 13 | 5 | 0 | 0 | 2.905 | 0.625 |
| | % | 14.286 | 61.905 | 23.810 | 0.000 | 0.000 | | |
| Spreadability | Persons (n) | 5 | 12 | 4 | 0 | 0 | 3.048 | 0.669 |
| | % | 23.810 | 57.143 | 19.048 | 0.000 | 0.000 | | |
| Absorbency | Persons (n) | 4 | 14 | 3 | 0 | 0 | 3.048 | 0.590 |
| | % | 19.048 | 66.667 | 14.286 | 0.000 | 0.000 | | |
| Fragrance | Persons (n) | 0 | 12 | 8 | 1 | 0 | 2.524 | 0.602 |
| | % | 0.000 | 57.143 | 38.095 | 4.762 | 0.000 | | |
| Overall usability | Persons (n) | 2 | 15 | 4 | 0 | 0 | 2.905 | 0.539 |
| | % | 9.524 | 71.429 | 19.048 | 0.000 | 0.000 | | |
| \* : Very Good (4), Good (3), Average (2), Poor (1), Very Poor (0) | | | | | | | | |

### Slide 16
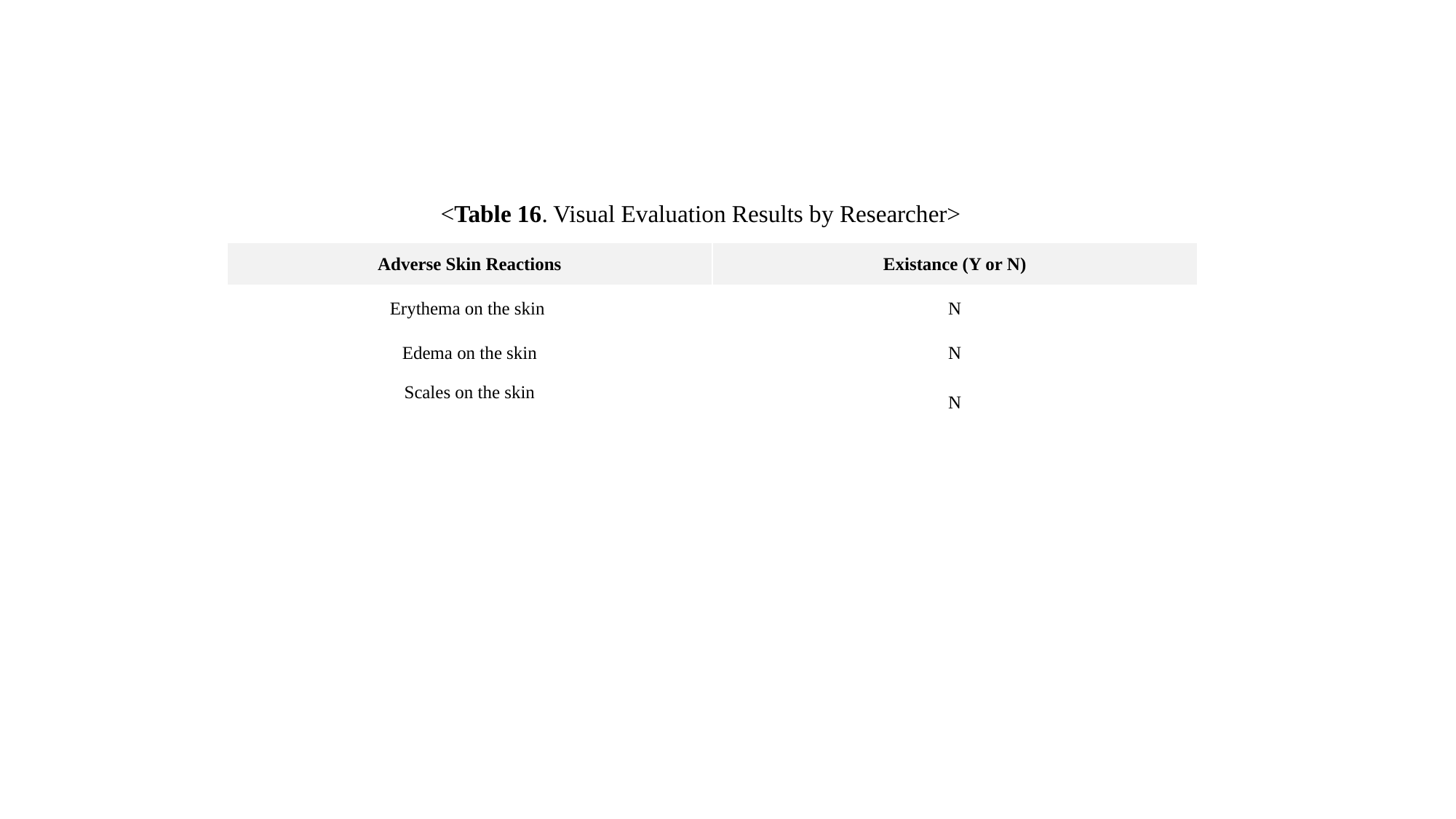

<Table 16. Visual Evaluation Results by Researcher>
| Adverse Skin Reactions | Existance (Y or N) |
| --- | --- |
| Erythema on the skin | N |
| Edema on the skin | N |
| Scales on the skin | N |

### Slide 17
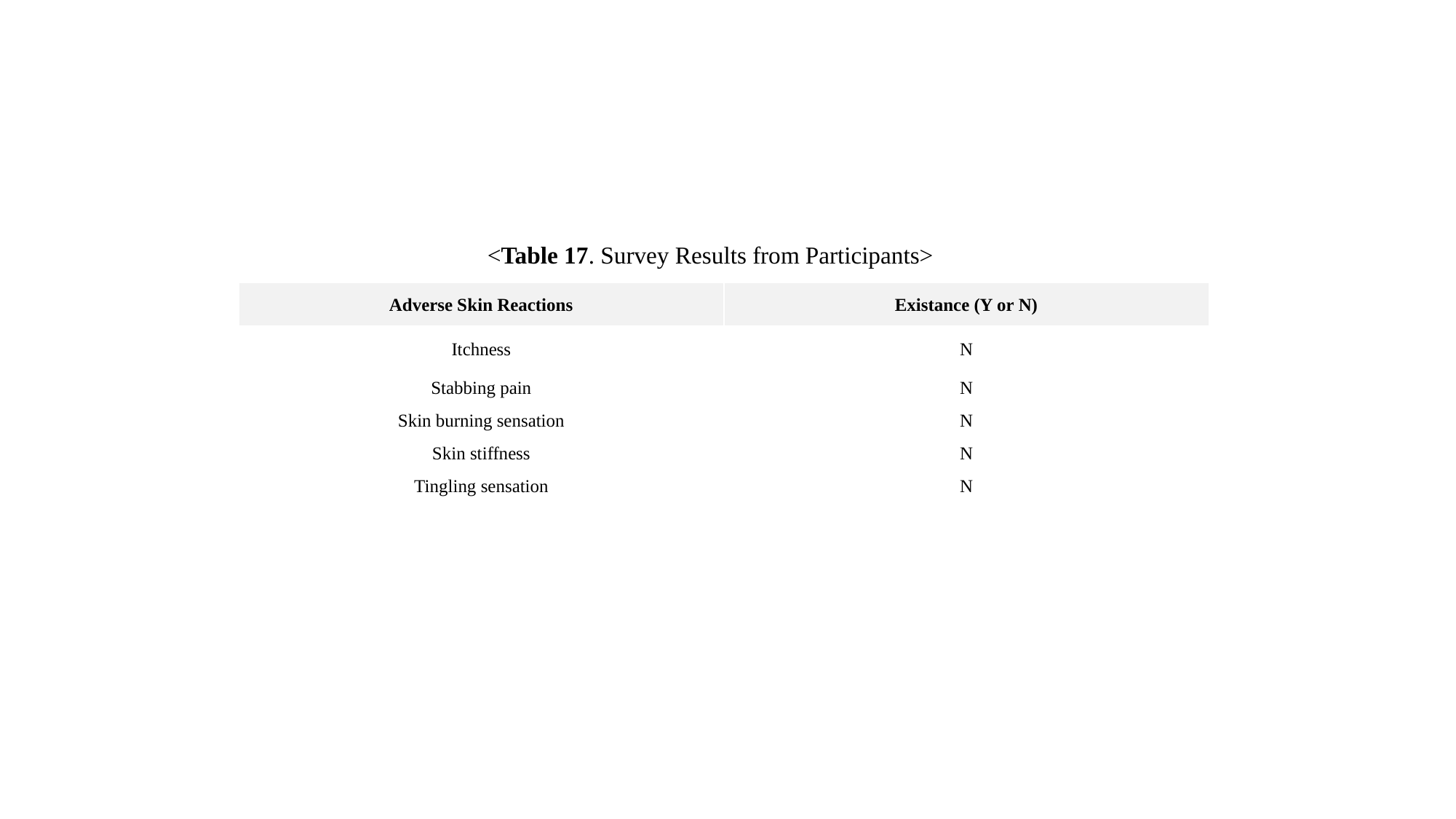

<Table 17. Survey Results from Participants>
| Adverse Skin Reactions | Existance (Y or N) |
| --- | --- |
| Itchness | N |
| Stabbing pain | N |
| Skin burning sensation | N |
| Skin stiffness | N |
| Tingling sensation | N |
